# Characterization of the mutational landscape of high-grade gliomas in a Latin American cohort

**DOI:** 10.1101/2023.12.05.23299374

**Authors:** Rodrigo Fernández-Gajardo, Hery Urra, Mauricio Saéz, Philippe Pihán, Beatriz Fonseca, Carolina Sanchez-Doñas, Maruf M. U. Ali, Sarah L. Rouse, Gabriel Cavada, Claudia Tissera, Rómulo Melo, David Rojas, José M. Matamala, Claudio Hetz

**Affiliations:** Biomedical Neuroscience Institute (BNI), Faculty of Medicine, University of Chile, Santiago, Chile; Department of Neurological Sciences, Faculty of Medicine, University of Chile, Santiago, Chile; Neurosurgery Department, Asenjo Neurosurgery Institute, Santiago, Chile; FONDAP Center for Geroscience, Brain Health, and Metabolism (GERO), Santiago, Chile; Program of Cellular and Molecular Biology, Institute of Biomedical Sciences, University of Chile, Santiago, Chile; Facultad de Odontología y Ciencias de la Rehabilitación, Universidad San Sebastián, Bellavista, Santiago, Chile; Genomics and Bioinformatics Center, Faculty of Medicine, Mayor University; Department of Life Sciences, Imperial College London, South Kensington, London SW7 2AZ, UK; Public Health Department, Faculty of medicine, University of Chile, Santiago, Chile; Pathology Department, Asenjo Neurosurgery Institute, Santiago, Chile; Buck Institute for Research on Aging, Novato, CA, 94945, USA

**Author notes:** **Address correspondence to:** José M. Matamala, Institute of Biomedical Sciences, University of Chile, Santiago, Chile., Claudio Hetz, Institute of Biomedical Sciences, University of Chile, Santiago, Chile. or. www.hetzlab.cl.

## Abstract

**Background:** Glioblastoma, an aggressive form of high-grade glioma, exhibits variations in the incidence and mortality patterns between populations of different ancestry. However, Hispanic and Latino populations remain largely underrepresented in studies of molecular characterization. Here we characterized the mutagenic landscape of high-grade gliomas within a Chilean population focusing on well characterized genetic variants associated with glioblastoma classification, progression, and prognosis.

**Methods:** We conducted a targeted genomic analysis using next-generation sequencing techniques in 70 Chilean patients with high-grade gliomas from a national single-center referral institution. We focused on the most relevant molecular markers such as isocitrate dehydrogenase 1/2 (IDH) mutations, telomerase reverse transcriptase promoter (TERTp) mutations, histone 3 (H3) gene family mutations, TP53 and PTEN mutations, epidermal growth factor receptor (EGFR) gene amplification, and cyclin-dependent kinase inhibitor 2A (CDKN2A) deletions. Survival analyses were performed to assess the clinical relevance of these markers and their impact on patient prognosis. Additionally, due to our interest in the role of proteostasis and glioblastoma, we investigated possible ERN1 variants in our Chilean cohort.

**Results:** Our findings mostly align with other international cohorts of non-Latin-American origin, confirming the importance of established molecular markers in glioblastoma. Notably, we identified novel TP53 and PTEN mutations with a predicted damaging effect, expanding the genetic spectrum of alterations in these brain tumors. Furthermore, a lower-than-expected mutation rate in the NF1 gene was observed, emphasizing a distinctive genetic profile of glioblastoma in this Latin American population. Prognostic assessments based on TERT promoter and IDH mutations mirrored previous studies on non-Latino American and European populations. Of note, we identified the germline ERN1 variant rs139229826 in 11% of our patient cohort, a variant previously unreported in glioblastoma patients.

**Conclusion:** This study underscores the importance of conducting glioblastoma research in underrepresented populations, providing insights into the molecular characteristics of high-grade gliomas within a Latin American context. Our findings contribute to the growing body of evidence suggesting molecular diversity across diverse glioblastoma populations, offering a foundation for future international comparative studies.

## Introduction

Glioblastoma is an aggressive and lethal brain cancer that constitutes a significant challenge in the field of oncology. A comprehensive understanding of the molecular mechanisms driving glioblastoma is crucial for the development of effective diagnostic and therapeutic approaches. While extensive research has been conducted on glioblastoma across diverse populations in the world (Barthel et al., 2019; Ceccarelli et al., 2016; Jonsson et al., 2019; Koo et al., 2020; Vaubel et al., 2020; L. B. Wang et al., 2021; Weinstein et al., 2013; J. Zhao et al., 2019); a notable knowledge gap exists concerning the incidence of mutations associated with glioblastoma in Latin-American populations. For example, in the largest cohort of patients with GBM and genomic data currently available – the TCGA pan-cancer atlas GBM database – only around 2% are individuals of Hispanic or Latino ethnicity (Weinstein et al., 2013). Thus, there is a pressing need to expand our current understanding of the spectrum of known glioblastoma mutations in the Latin-American population.

With the increasing integration of molecular criteria into the widely used WHO classification of tumors of the central nervous system (CNS), the importance of accurate and reliable molecular techniques has also increased in clinical practice (Sahm et al., 2023). The current criteria for diffuse high-grade astrocytoma classification include disease-defining alterations such as isocitrate dehydrogenase 1/2 (IDH) mutations and histone 3 (H3) gene family (H3F3A, H3C2) mutations, as well as grade-defining alterations such as telomerase reverse transcriptase promoter (TERTp) mutations, epidermal growth factor receptor (EGFR) gene amplification and the combined gain of entire chromosome 7 and loss of entire chromosome 10 in the case of IDH-wildtype tumors, or cyclin-dependent kinase inhibitor 2A/B (CDKN2A/B) deletion in the case of IDH-mutant tumors (Louis et al., 2021). Molecular classification profoundly influences the prognosis and survival of glioblastoma patients, directly impacting their treatment, follow-up, and care (Berger et al., 2022; Gritsch et al., 2022; Horbinski et al., 2022; Kurokawa et al., 2022).

Molecular diagnostics using Next-Generation-Sequencing (NGS)-based methods have become the gold-standard for multiple diagnostic workups (Sahm et al., 2023). The current study aims to investigate the mutational landscape of glioblastoma on a Latin-American cohort, focusing on Chile as a nation characterized by its diverse ethnic composition, including Indigenous, European, and African influences (Eyheramendy et al., 2015). Our research not only seeks to provide insight into the genetic drivers of glioblastoma in this specific south American population, but also underscores the broader importance of studying ethnic-specific genetic variations and their potential impact on tumor biology and clinical outcomes.

Using an NGS-based approach, we describe the frequency of some of the most common glioblastoma mutations in a Chilean cohort. Importantly, we found that these mutations exhibit similar prognostic significance compared to the global population, reinforcing their importance as key drivers of glioblastoma progression. However, our study also unveils a unique aspect of glioblastoma genetics in Latin-America. We identified the presence of a likely germline single-nucleotide polymorphism (SNP) previously reported in healthy Latin American populations, and established its elevated prevalence in our glioblastoma cohort. Furthermore, using structural bioinformatic approaches we predicted that this specific SNP may exert a functional influence on inositol-requiring enzyme 1 α (IRE1α), the protein encoded by *ERN1*, a critical player in cellular stress responses to proteostasis alterations (Hetz et al., 2020). This discovery not only underscores the necessity of ethnically tailored tumor characterization but also calls for comprehensive investigations into the potential function of this IRE1α variant in the biology of glioblastoma.

## Methods

### Patient selection

We conducted an observational retrospective cohort study, which included 70 patients aged 18 and above with a confirmed diagnosis of high-grade glioma based on the current WHO classification system for CNS tumors at the time. Patients who underwent total or partial resection surgery at the Asenjo Neurosurgery Institute between January 2014 and January 2020 were included in the study. Patients with previous diagnosis of diffuse glioma, incomplete follow-up, insufficient tumor tissue samples for molecular study, and patients with known hereditary neoplastic syndromes were excluded. No other exclusion criteria were applied. Additionally, 5 patients who underwent anterior temporal lobectomy for epilepsy treatment were prospectively included in the analysis as non-tumor tissue controls, and specific informed consents were obtained. The study was approved by the ethics committee of the Metropolitan East Health Service. Funding was secured through competitive grants from the Metropolitan East Health Service and the Department of Neurological Sciences of the University of Chile.

### Panel Design, Parallel Mass Sequencing and Bioinformatic Analysis

Formalin-fixed paraffin-embedded tissue samples were obtained from the Pathology Service at the Asenjo Neurosurgery Institute. After neuropathologist confirmation of a tumor percentage greater than 70% in each area to be extracted, 3-6 sections of 10 microns were prepared from each paraffin block using the GeneJET FFPE DNA Purification Kit. No matched blood samples were available. The extracted DNA was quantified using fluorescence-based methods (Quant-iT™ PicoGreen™ dsDNA and/or Qubit™ dsDNA HS), resulting in an average concentration of 40 ng/µL. We utilized the AmpliSeq™ for Illumina tool to create a customized amplicon panel covering specific genomic regions of interest of several neuro-oncology related genes. Databases such as cbioportal, NCBI/Clinvar, and the Catalogue of Somatic Mutations in Cancer (COSMIC) were employed to determine the relevance and genomic coordinates of sites of interest. Amplicons were designed to encompass these mutations (Supplementary Table 1). This sequencing strategy included 339 amplicons with an average length of 134 base pairs (ranging from 125 to 140 base pairs) and covered 15,596 base pairs, including the following genes: IDH1, IDH2, H3F3A, H3C2, EGFR, CDKN2A, ATRX, SMARCAL1, BRAF, TP53, PTEN, and ERN1. Because the promoter region of the TERT gene contains a high percentage of GC bases (Chiba et al., 2015), a complementary sequencing strategy was designed to cover the region using 25 amplicons with an average length of 230 base pairs (ranging from 192 to 266 base pairs) that covered 5,750 base pairs. 1 amplicon was designed to cover the TERT promoter region, and the rest of the amplicons were designed to either give additional coverage for the most relevant hotspots or to assess additional regions of interest (Supplementary Table 1). To reduce the occurrence of false negatives in sequencing, a coverage depth of greater than 200 reads per position of interest was planned to detect allelic frequencies as low as 5%, with a threshold of ≥10 altered reads, following literature recommendations (Petrackova et al., 2019).

Library preparation and parallel mass sequencing were conducted at the Genomics and Bioinformatics Center of the Mayor University. An AmpliSeq™ Custom DNA Panel kit and a TruSeq™ nano DNA kit from Illumina were used to prepare libraries for each sequencing strategy, respectively. For both AmpliSeq™ and TruSeq™ libraries, library generation efficiency was evaluated with different initial amounts of DNA to adjust the various stages of the process based on the specific sample and panel characteristics. Automated electrophoresis of the first libraries were performed for quality control and size estimation using the DNA 1000 kit on the Agilent 2100 Bioanalyzer system. All libraries were quantified by qPCR using the Illumina KAPA library quantification kit and were pooled at a concentration of 10 nM before sequencing. A Mid Output Kit v2.5 (150 cycles) was used on a NextSeq sequencer with 25% phi-X for all Ampliseq™ libraries, and a Micro Kit v2 (300 cycles) with 25% phi-X on a MiSeq sequencer was used for all TruSeq™ libraries.

*.fastQ files were generated from the raw sequencing data and analyzed using the fastQC software for quality control (https://www.bioinformatics.babraham.ac.uk/projects/fastqc/). Reads of low quality and adapter contamination were removed using the trim_galore tool (Bolger et al., 2014). Processed reads were aligned to the GRCh38 reference genome using the Bowtie2 tool (Langmead & Salzberg, 2012). The aligned *.sam results were then converted to *.bam files using the samtools application for subsequent analysis. Somatic mutations were called according to GATK recommendations (Depristo et al., 2011; McKenna et al., 2010; Van der Auwera et al., 2013), including additional considerations for amplicon panels, as described below. Bam files were analyzed with MarkDuplicatesSpark and BaseRecalibrator, using gnomadV2 database (Gudmundsson et al., 2022), to finally recalibrate with ApplyBQSR to meet GATK standards. Somatic variants were detected using Mutect2 (Benjamin et al., 2019), only on target regions. To identify and discard artifactual variants, GetPileupSummaries with CalculateContamination, LearnReadsOrientation and a panel of normal were used, Additionally, the Integrative Genomics Viewer (IGV 2.10) was used to identify and correct potential errors. Finally, we used FilterMutectCalls to recover clustered events and to discard variants with an allelic frequency <0.05 or <10 reads supporting the variant.

Variants in vcf files were annotated with Funcotator and reports were generated in vcf and maf files (Depristo et al., 2011). All genomic coordinates are mapped to the GRCh 38 assembly. Posterior analysis was performed with Variant Effect Predictor (McLaren et al., 2016) to predict pathogenicity. Novel variants and variants not previously reported in gliomas that are predicted to result in single amino acid substitutions were subsequently evaluated using Sorting Intolerant From tolerant (SIFT) and polymorphism phenotyping-2 (PolyPhen-2) to predict the effect of the change on protein function (Adzhubei et al., 2010; Kumar et al., 2009) while variants that resulted in frameshifts were evaluated using SIFT indel (Hu & Ng, 2012). Maftools R package was used to prepare oncoplots (Mayakonda et al., 2018).

Copy number analysis of EGFR, CDKN2A and PTEN was performed using the R package CNVPanelizer (Oliveira & Wolf, 2020), which normalizes the number of reads for the gene of interest based on the total reads for each sample, followed by a resampling algorithm (bootstrapping with replacement, n=10,000) to mitigate the effect of coverage heterogeneity in data interpretation. The fold change of EGFR, CDKN2A and PTEN in relation to 5 control samples were determined, using coverage information from 136 EGFR amplicons, 21 CDKN2A amplicons, 46 PTEN amplicons and 120 amplicons outside the regions of interest (ATRX, NF1, BRAF, and TP53). The statistical significance of the results was assessed using the Bonferroni test, and only CNVs that were significant are reported.

### 3D modelling, structural and dynamic protein analyses

As we explored the effect of the ERN1 variant rs139229826, the resulting IRE1 L914M mutant was generated *in silico* by performing a single amino acid mutation substitution from Lysine to Methionine using the human IRE1 cytosolic region coordinate file (PDB 3P23). The subsequent IRE1 mutant structure model was displayed in cartoon representation and visualised in pymol (Schrodinger Inc) with the images ray-traced to enhance picture quality. Dynamic models showing solution atomized molecular simulations of IRE1 wildtype and IRE1 L914M (based on a point mutation generated *in silico*) were made using 200-500ns simulation times under a Charmm36m forcefield, employing a 150mM NaCl solution.

### Statistical analysis

Associations between categorical variables were assessed by the χ2 test. Kaplan–Meier estimates and log-rank testing were performed for survival curves analyses. Proportional hazards Cox regressions were used to examine the relationship between predictor variables and overall survival, and a stepwise forward selection was used when multiple predictors were analyzed.

### Results and Discussion

#### Clinical outcomes

Our studied cohort included 70 patients diagnosed with high-grade astrocytomas. The most relevant clinical variables measured, including access to different treatments and outcomes for all patients, are presented in **table 1**. Patients exhibited an average age of 57.28 years with a significant majority (96%) demonstrating grade IV astrocytomas. All patients in this cohort underwent surgical resection as part of their treatment plan; however, mainly due to financial limitations, only 20% received a complete regimen of adjuvant therapy with radiation and temozolomide. Patients who received the complete Stupp protocol were younger and presented a more favorable postoperative Karnofsky Performance Status (KPS). These patients had a considerably longer median survival time, with an average survival of 28.7 months compared to 5.8 months of those who only received surgery with or without adjuvant radiotherapy.

**Table 1.**
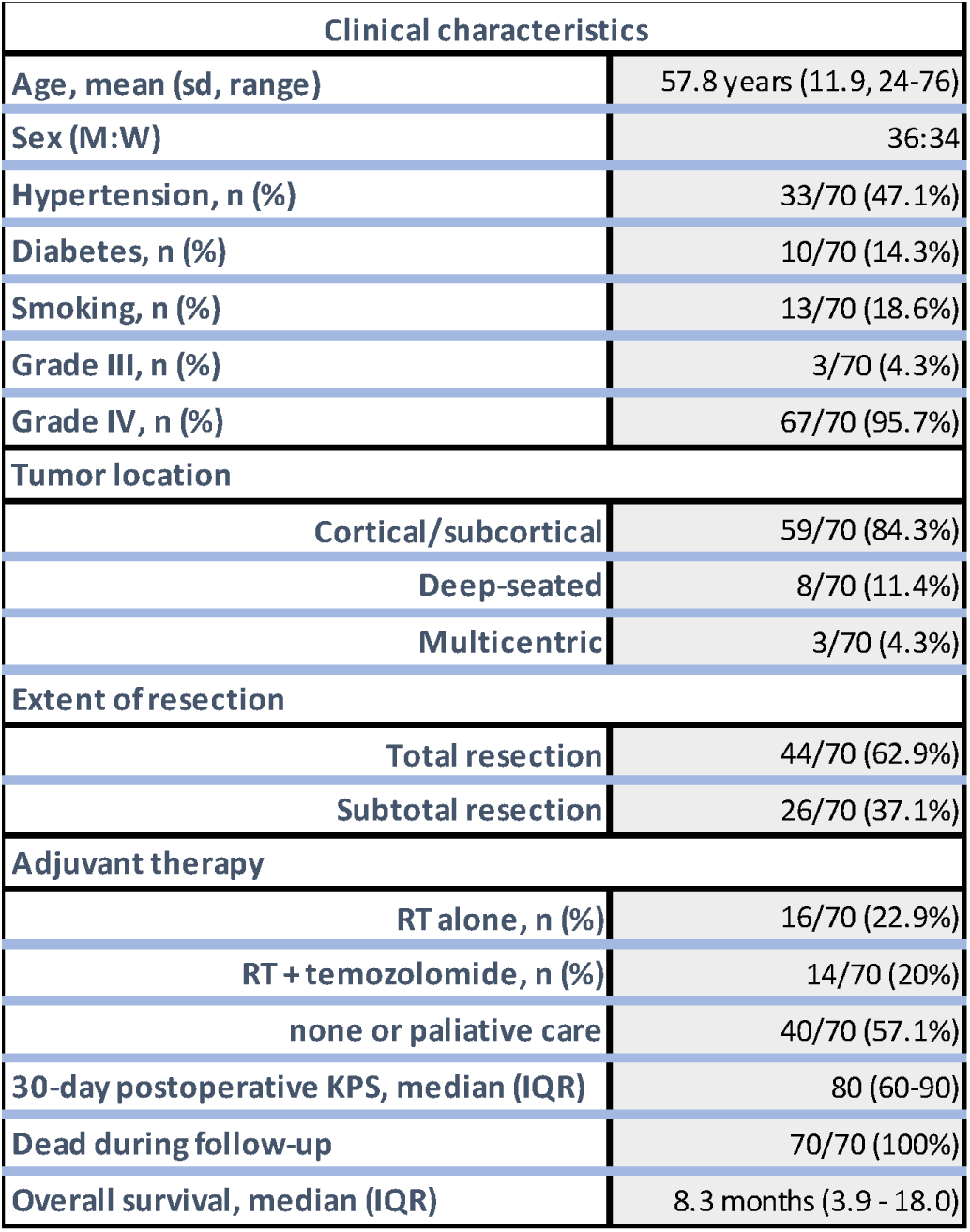
Baseline characteristics of 70 high-grade glioma patients. Demographics, comorbities, tumor characteristics, therapies and clinical outcomes are shown. RT = radiotherapy, KPS = Karnosfsky performance score, IQR = Interquartile range.

#### Sequencing metrics

A Q-score greater than Q30 was calculated for over 90% of the sequencing reads and were subsequently used for variant calling. The percentage of reads on-target exceeded 90% in all patients. 61 unique variants were identified in 66 out of 70 patients (**supplementary table 2 and Figure 1**). 67 out of 70 patients had a genetic alteration when considering CNVs. The sequencing depths over each variant are shown in the **supplementary table 3** for all samples.

**Figure 1.**
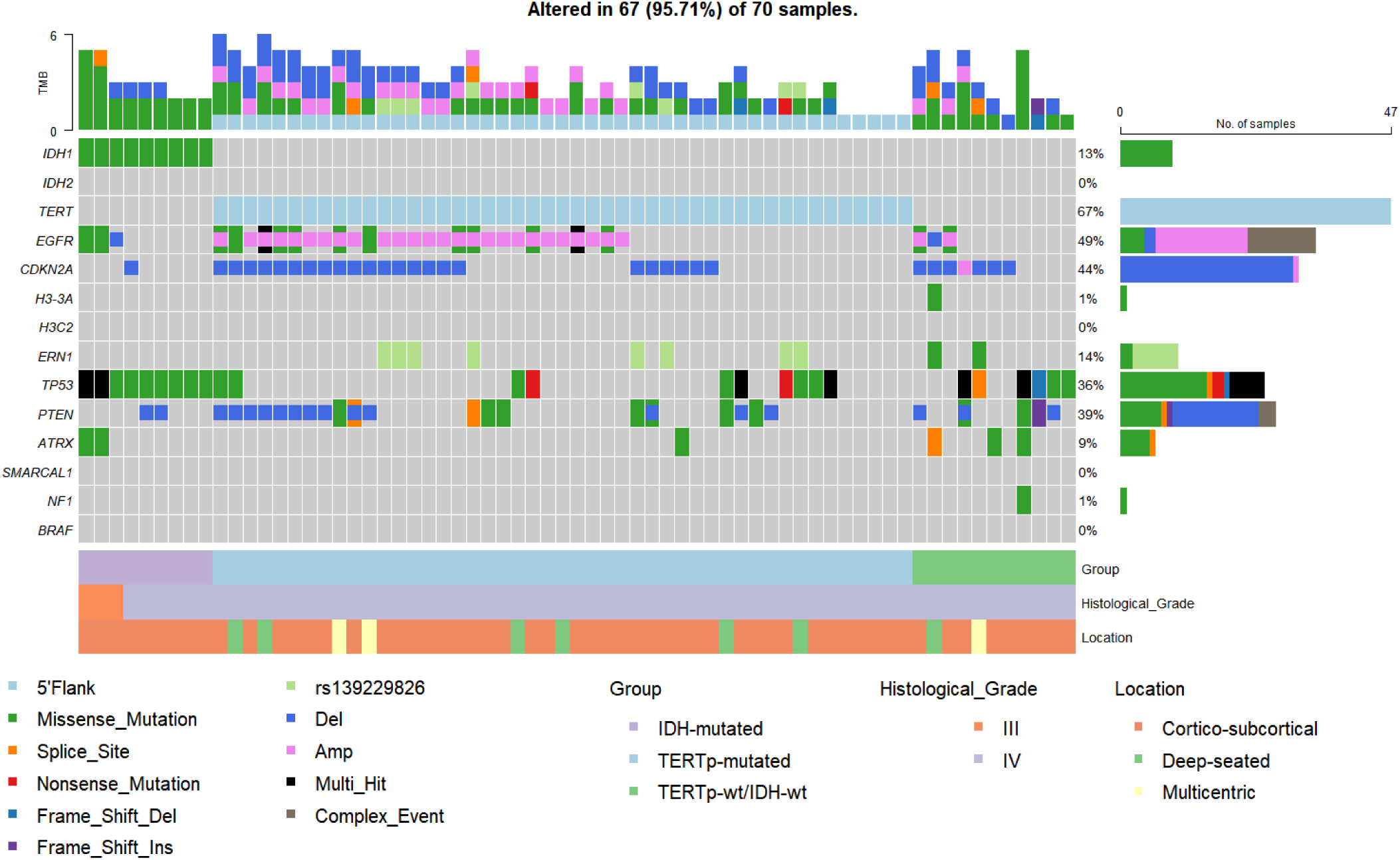
Co-mutation plot displaying genetic alterations in 15 genes included in our sequencing panel. Nex-generation sequencing results in 70 patients with high-grade glioma are shown. Groups with different clinical outcomes, histological grade and tumor location for each patient are displayed below. All the multi-hit alterations in TERT promoter gene included the canonical mutations 113 and 135. Multi-hit = 2 or more variants were identified; Complex event = A copy number variation and another genomic alteration occurred simultaneously; Amp = Gene amplification; Del = Gene deletion; IDH, Isocitrate Dehydrogenase; TERTp, Telomerase Reverse Transcriptase Promoter. TMB = Tumor mutational burden.

#### Frequency of the most common genomic alterations in glioblastoma

Next, we characterized the mutational status of 70 patients from a Latin-American population that included several brain cancer-related genes used in molecular classification (IDH1, IDH2, H3F3A, H3C2, EGFR, CDKN2A, alpha-thalassemia/mental retardation, X-linked (ATRX), SWI/SNF Related, Matrix Associated, Actin Dependent Regulator Of Chromatin, Subfamily A Like 1 (SMARCAL1), Serine/threonine-protein kinase B-raf (BRAF), tumor protein p53 (TP53), and Phosphatase and tensin homolog (PTEN)). Additionally, we included an additional gene that we are currently studying, Endoplasmic Reticulum to Nucleus Signaling 1 (ERN1) that encodes for IRE1α (Lhomond et al., 2018). A graphic representation of the genomic alterations identified in this cohort are presented in **Figure 1**. Our findings and comparisons to other populations for specific genes are detailed in the following paragraphs.

#### Mutations included in the WHO classification of CNS tumors

IDH is arguably the most relevant genetic marker currently in use for the classification of astrocytic tumors and when mutated, defines the recently termed *IDH-mutant astrocytoma*, which can be categorized as grade II, III or IV depending on additional histological and/or molecular criteria. This differentiation distinguishes IDH-mutated tumors from glioblastomas, which requires an IDH-wildtype confirmation. In our patient cohort, we found that ∼13 % of patients had IDH1 mutations (9 out of 70): 8 had the most frequent R132H mutation, while 1 had the less common R132S mutation. Clinical characteristics of these patients were similar to the characteristics reported in other cohorts (Killela et al., 2014; Yan et al., 2009), significantly affecting younger subjects than those carrying IDH-wildtype alleles (49 years old (range 30 to 76) vs 59 years old (range 24 to 75)). When the subgroup of tumors histologically graded as IV was assessed, the frequency of IDH1 mutations was 8.5% (6 out of 67), which is comparable to other cohorts (Ceccarelli et al., 2016; Cerami et al., 2012; Gao et al., 2013). Consistently with their much lower frequency (Yan et al., 2009), no IDH2 mutations were identified in our cohort. Also, in agreement with previous reports (Killela et al., 2014), none of the patients with IDH1 mutations exhibited concomitant TERTp mutations.

Immunohistochemistry (IHC) for IDH1-R132H was routinely performed in patients diagnosed after 2017 (n = 38) and it showed a perfect correlation with the sequencing results, with all 6 cases that had a positive R132 IHC showing a corresponding IDH1 mutation. Interestingly, although the mean age of the 9 patients carrying IDH1 mutations was 49 years old, two of these patients were older than 55 years old, which is the most common cut-off for recommending sequencing in case of negative R132H IHC (Louis et al., 2021; Nassiri et al., 2017). Moreover, the patient that carried the R132S mutation was older than 55 years-old and could not have been accurately diagnosed using IHC, which was expected since immunodetection is exclusively directed at the R132H protein. This technical limitation highlights the relevance of using genomic techniques to evaluate IDH1 mutations in glioma patients.

Less common mutations that also define specific entities according to the current diagnostic criteria are those in the H3 gene family, including H3F3A and HIST1H3B genes (Castel et al., 2015). In our cohort, we identified only one patient with H3 mutations—a subject aged between 26 and 30 years old, presenting with a thalamic tumor that exhibited the most frequent H3F3A mutation (K28M, rs1057519903), consistent with a different entity, a *diffuse midline glioma, H3 K27-altered*. No H3F3A G35 nor any HIST1H3B alterations were found in our patients. Consistent with international cohorts (Schwartzentruber et al., 2012; Sturm et al., 2012; Wu et al., 2012), our findings indicate that adult patients with tumors located in different supratentorial regions are anticipated to demonstrate a low frequency of H3 gene mutations.

In addition to the entity-defining markers, we evaluated the frequency of three genomic alterations that are currently considered in the grading of diffuse astrocytomas: TERTp mutations and EGFR amplification (molecular criteria for a grade IV glioblastoma) and CDKN2A deletion (molecular criterion for a grade IV IDH-mutant astrocytoma). TERTp mutations typically occur in 70-80% of primary glioblastoma cases (Arita et al., 2013; Killela et al., 2013). These mutations are closely associated with an increased TERT gene expression, resulting in the preservation of telomeres playing a pivotal role in promoting gliomagenesis (Bell et al., 2015; Ceccarelli et al., 2016). We evaluated the two canonical TERTp hotspots where mutations normally occur (Nonoguchi et al., 2013). We found that 47 out of 70 patients (67.1%) presented one of these mutations, with 37 patients presenting TERTp mutations at position g.chr5:1295113G>A and 10 patients presenting the less frequent g.chr5:1295135G>A mutation. These results are consistent with previous work in European, North American, and Asian populations (Labussière, Di Stefano, et al., 2014; Nguyen et al., 2017; Simon et al., 2015; Wu et al., 2012), and we will further discuss their clinical relevance in the following sections.

Amplification of the epidermal growth factor receptor (EGFR) is a recognized molecular hallmark characterizing high-grade gliomas. Notably, the EGFR gene has been the focal point of preliminary clinical trials, which exhibited promising initial prospects but ultimately failed to yield favorable outcomes (Lee et al., 2020). Mechanistically, EGFR amplification is an early event in glioma progression (Del Vecchio et al., 2013), which results in its persistent activation, promoting tumor growth and invasion (Talasila et al., 2013). In our cohort, we found that 28 patients (40%) exhibited a significant increase in EGFR gene copy numbers. EGFR amplification was present only in IDH-wildtype tumors, and it was more frequent in TERTp-mutated cases (55.3% vs 14.3%; p<0.01). On the other hand, CDKN2A deletion plays a critical role in regulating cell cycle and has increasingly been recognized as a prognostic marker in IDH-mutant astrocytomas (Appay et al., 2019; Lu et al., 2020a, 2020b). CDKN2A partial or total deletions were present in 30 patients (42.9%). Only one patient with a CDKN2A deletion also carried an IDH1 mutation. These frequency distributions and their co-existence with TERTp and IDH mutations are consistent with other brain cancer cohorts (Brennan et al., 2013).

Regarding effective grading changes based on molecular markers, all patients with IDH-wildtype tumors that also carried TERTp mutations and/or EGFR amplification, as well as patients with IDH-mutant tumors that also carried CDKN2A deletions were considered grade IV tumors based on histological features, therefore no grading changes were made based on these findings.

#### Additional mutations related to glioblastoma progression

TP53 and PTEN genomic alterations are among the most prevalent mutations in gliomas, and multiple targeted therapies focused on these genes have been evaluated over the last few years (Hernández Borrero & El-Deiry, 2021; Hoxhaj & Manning, 2019; H. fu Zhao et al., 2017). We sequenced the regions containing all the previously described mutations in TP53 and PTEN in glioblastomas and found a similar mutation rate for both genes, compared to other cohorts (Cerami et al., 2012; Gao et al., 2013). Specifically, 25 out of 70 patients (allelic frequency (AF): 35.7%) carried a TP53 mutation, 3 of which had not been previously reported in glioma patients (g.chr17:7669615delT (AF: 45% SIFT indel: neutral effect), g.chr17:7674926C>T (rs587778719; AF: 10.6%; SIFT: tolerated; Polyphen-2: benign), and g.chr17:7674966delC (AF: 84.4%; SIFT indel: damaging). On the other hand, PTEN alterations were found in 29 out of 70 patients (41.4%), with partial or total deletions present in 18 patients, while single nucleotide variants were present in 13 patients. Out of 11 unique PTEN variants identified, two corresponded to novel variants in glioma patients (g.chr10:87925553A>G (AF: 43.6%; SIFT: deleterious; Polyphen-2: probably damaging) and g.chr10:87933224_87933225insG (AF: 43.7% SIFT indel: damaging). Thus, novel PTEN (frameshift deletion g.chr17:7674966delC;) and TP53 variants (missense mutation g.chr10:87925553A>G and frameshift insertion g.chr10:87933224_87933225insG) with a predicted damaging effect were identified in our study, expanding the mutational landscape of these genes in glioma patients. However, each of these variants was only present in one patient, and 4 out of 5 carried an allelic frequency that suggests a somatic origin, therefore it is unlikely that they constitute a specific hallmark of the Latin-American population.

ATRX and SMARCAL1 mutations are linked to telomerase-independent telomere maintenance in glioblastoma (Brosnan-Cashman et al., 2021; Diplas et al., 2018; Liu et al., 2023; Nandakumar et al., 2017; Qin et al., 2022). While ATRX mutations are typically present in about 20% of high-grade gliomas (Haase et al., 2018), our study identified a lower-than-expected frequency corresponding to 10% (7 out of 70), and none of the most reported alterations were present in our cohort. However, numerous unique ATRX variants not covered by our sequencing design have been described. Unexpectedly, one patient with an ATRX mutation also presented a TERTp mutation in our cohort (**Fig. 1**). We screened the most common SMARCAL1 cancer-associated mutations in our sequencing panel, including Arg645Ser (R645S), Phe793del (del793), and Gly945fs*1 (945 fs) mutations, which have been previously described in up to 20% of TERTp-wt/IDH-wt tumors (Diplas et al., 2018). In contrast, no SMARCAL1 mutations were found in our cohort.

Regarding NF1, previous cohorts of glioblastoma (TCGA pancancer atlas GBM database, MSKCC dataset) have reported approximately a 13% mutation rate for this gene (Cerami et al., 2012; Gao et al., 2013; Jonsson et al., 2019). Interestingly, although our panel design covered most of the common reported variants, none of these variants were present in our cohort, suggesting that NF1 mutations are less frequent compared to other populations and/or located in different genomic regions. Instead, we identified one variant of uncertain significance (rs2151559381) present in 1 patient, that has not been previously reported in glioma patients.

Finally, we also examined the BRAF gene for the classic V600E mutation, traditionally associated with epithelioid features of glioblastoma (Di Nunno et al., 2022; McNulty et al., 2021). Despite adequate coverage, our sequencing revealed no BRAF mutations in our cohort.

#### Survival Analyses – Association with relevant genes and clinical data

Next, we compared overall survival curves between patient groups defined by the presence of IDH mutations, TERTp mutations or the combination of both, as the prognostic significance of these markers has been well-documented (Labussière, Boisselier, et al., 2014; Labussière, Di Stefano, et al., 2014; Simon et al., 2015). In our study, IDH mutations were associated to significantly longer overall survivals than IDH-wildtype patients (32.8 vs 7.2 months; p < 0.01; Fig. 2A), while patients with TERTp mutations had lower overall survival compared to those without TERTp mutations (6.4 vs. 16.3 months; p < 0.01; **Figure 3A**). Our analysis revealed that EGFR, TP53, PTEN, or CDKN2A were not associated with a significant difference in overall survival (not shown), a description that is consistent with previous reports (Kraus et al., 2000; Marker & Pearce, 2020). Moreover, comparing survival curves based on both TERTp and IDH mutations resulted in further discrimination of patient groups with different survival (32.8 vs 10.5 vs 6.4 months; mantel-cox and logrank test for trend *p* < 0.01), emphasizing the additive role of these mutations in predicting patient survival outcomes.

**Figure 2.**
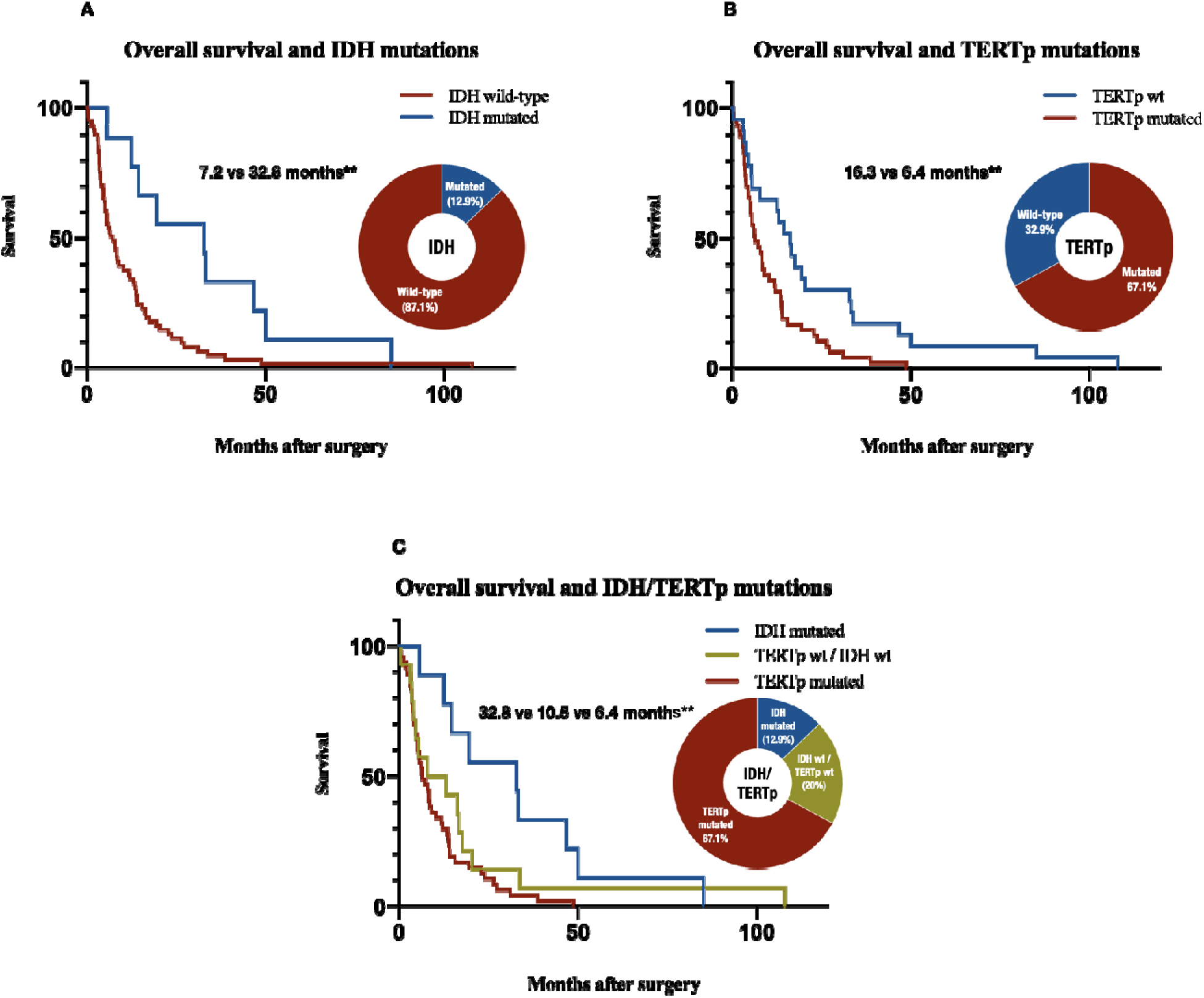
Stratified overall survival curves. Stratified Kaplan-Meier curves and distribution of mutation status are presented for (A) IDH mutations, (B) TERTp mutations and (C) a combined classification using IDH and TERTp mutations. Log-rank test was performed to compare curves, and double asterisks indicate *p*-values < 0.01. IDH, Isocitrate Dehydrogenase; TERTp, telomerase reverse transcriptase promoter.

**Figure 3.**
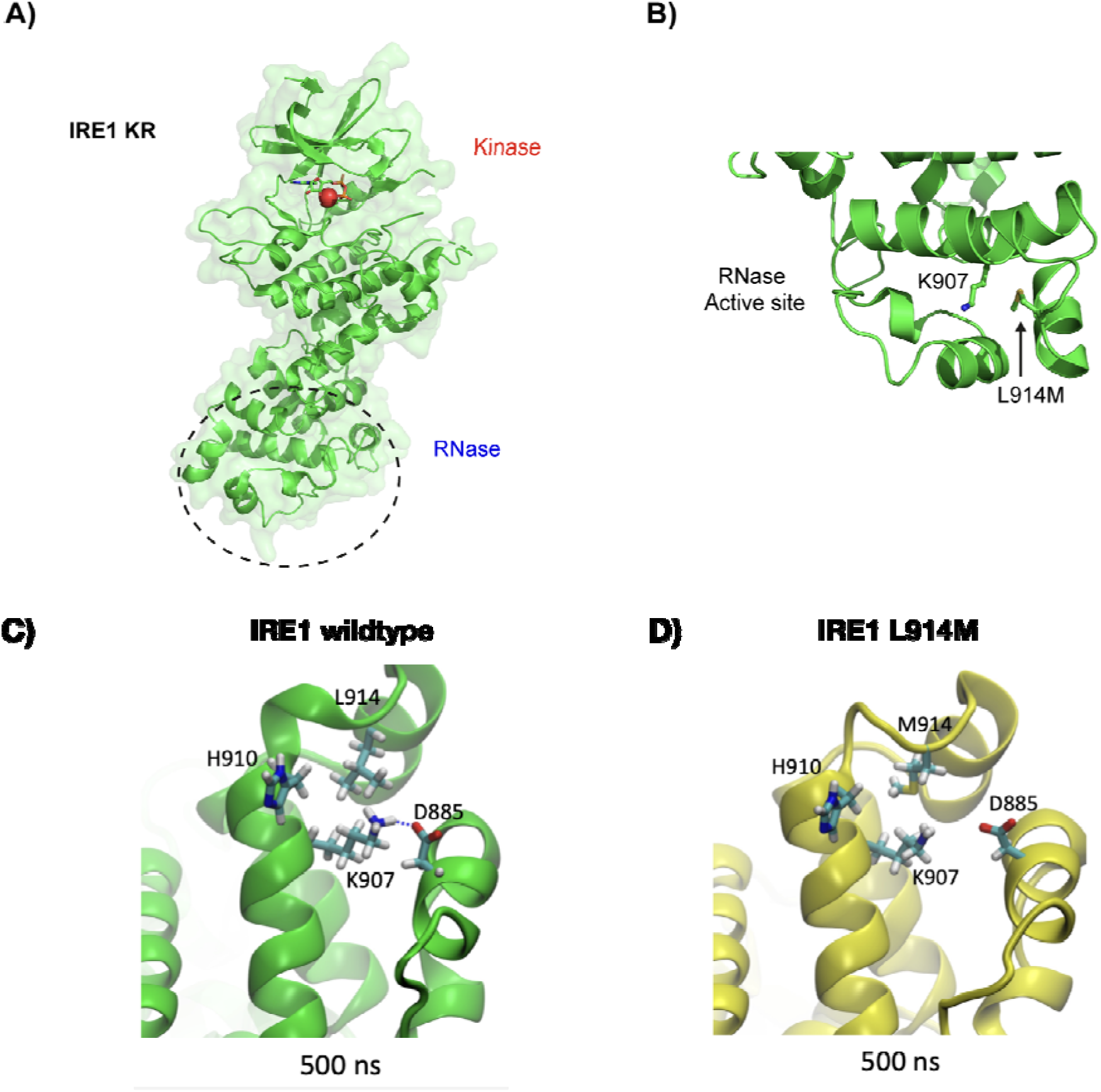
Structural and dynamic modelling of IRE1 mutant. A) Overview of human IRE1 cytosolic region structure consisting of kinase and RNase domains (PDB: 3P23). B) A close-up view of RNase domain active site highlighting the key catalytic Lys907 residue and the Met914 mutant. Even though the L914M mutation is in close proximity to the Lys907, it maintains the hydrophobic nature of the amino acid suggesting it will be structurally well tolerated by the protein. Dynamic models showing solution atomized molecular simulations of IRE1 wildtype (C) and IRE1 L914M (D) after 500ns under a Charmm36m forcefield are displayed. When comparing both models, we found that the ionic bond between K907 and D885 is lost in the mutated protein, suggesting that the L914M substitution is highly likely to modify IRE1 RNase activity.

A multivariable regression analysis was also conducted to evaluate the prognostic value of different clinical and molecular determinants in our cohort. The following factors were included in our analyses: sex, age, hypertension, diabetes mellitus, smoking, extent of tumor resection (total versus subtotal), 30-day postoperative KPS (favorable outcomes were defined as a score of 90 or 100), histological grade, radio- and chemotherapy (for patients with OS longer than 2 months), IDH mutations, TERTp mutations, EGFR amplification, CDKN2A deletion and the presence of the ERN1 variant rs139229826. We included all variables with univariate *p*-values < 0.25 in a multivariate analysis. The coefficients, confidence intervals, and significance values for univariate and multivariate analyses of each prognostic factor are shown in **Table 2**.

**Table 2.** Univariate and multivariate Cox regression models for clinical and genetic variables with a likely prognostic value for overall survival in high-grade glioma patients (n = 70). KPS, Karnofsky Performance Score; TERT, Telomerase Reverse Transcriptase; CDKN2A, Cyclin-Dependent Kinase Inhibitor 2A; IDH, Isocitrate Dehydrogenase; EGFR, Epidermal Growth Factor Receptor; ERN1, Endoplasmic Reticulum to Nucleus Signaling 1. † = multicentric or deep-seated location versus cortico-subcortical location. * = *p*-values < 0.25 kept for multivariate analyses. ** = *p*-values < 0.05 in multivariate analyses.

| Predictive variable | Univariate |  |  |  |  |  | Multivariate |  |  |  |  |  |
| --- | --- | --- | --- | --- | --- | --- | --- | --- | --- | --- | --- | --- |
|  | HR | Std. Err | z | p value | [95% CI] |  | HR | Std. Err | z | p value | [95% CI] |  |
| Sex | 0.93 | 0.23 | -0.29 | 0.77 | 0.58 | 1.50 |  |  |  |  |  |  |
| Age | 1.02 | 0.01 | 2.16 | **0.03\*** | 1.00 | 1.04 | 1.02 | 0.01 | 2.02 | **0.04\*\*** | 1.00 | 1.05 |
| Hypertension | 1.48 | 0.37 | 1.58 | **0.12\*** | 0.91 | 2.42 |  |  |  | 0.72 |  |  |
| Diabetes | 0.99 | 0.34 | -0.03 | 0.98 | 0.50 | 1.94 |  |  |  |  |  |  |
| Smoking | 1.21 | 0.38 | 0.61 | 0.54 | 0.65 | 2.24 |  |  |  |  |  |  |
| Tumor location† | 1.54 | 0.39 | 1.71 | **0.09\*** | 0.94 | 2.52 |  |  |  | 0.71 |  |  |
| Favourable postoperative KPS (90-100) | 0.48 | 0.13 | -2.61 | **<0.01\*** | 0.28 | 0.83 |  |  |  | 0.72 |  |  |
| Total vs subtotal resection | 0.35 | 0.09 | -3.91 | **<0.01\*** | 0.21 | 0.60 | 0.53 | 0.16 | -2.11 | **0.04\*\*** | 0.30 | 0.96 |
| Histological grade | 2.85 | 1.71 | 1.75 | **0.08\*** | 0.88 | 9.23 |  |  |  | 0.31 |  |  |
| Radiotherapy | 0.25 | 0.07 | -4.83 | **<0.01\*** | 0.14 | 0.44 | 0.34 | 0.12 | -3.15 | **<0.01\*\*** | 0.18 | 0.67 |
| Chemotherapy | 0.22 | 0.08 | -4.12 | **<0.01\*** | 0.11 | 0.46 | 0.29 | 0.13 | -2.70 | **<0.01\*\*** | 0.12 | 0.71 |
| IDH1 mutation | 0.39 | 0.14 | -2.58 | **0.01\*** | 0.19 | 0.79 | 0.27 | 0.11 | -3.09 | **<0.01\*\*** | 0.12 | 0.62 |
| TERT promoter mutation | 2.15 | 0.59 | 2.77 | **<0.01\*** | 1.25 | 3.69 |  |  |  | 0.19 |  |  |
| EGFR amplification | 0.98 | 0.25 | -0.08 | 0.94 | 0.60 | 1.60 |  |  |  |  |  |  |
| ENR1 variant | 1.28 | 0.49 | 0.64 | 0.52 | 0.61 | 2.69 |  |  |  |  |  |  |
| CDKN2A | 1.17 | 0.29 | 0.62 | 0.53 | 0.72 | 1.90 |  |  |  |  |  |  |

The final multivariable regression model identified five factors capable of explaining overall survival in our high-grade glioma patient series, including (i) therapeutic alternatives (total versus subtotal resection, and radiotherapy and/or chemotherapy access), (ii) patients’ age and (iii) IDH mutation status. It is noteworthy that although TERTp mutations exhibited considerable prognostic value in univariate analysis, they displayed significant interactions with other prognostic factors such as age, postoperative Karnofsky Performance Status (KPS), and the utilization of radio- and chemotherapy (p < 0.01). These interactions could elucidate the lack of significance when jointly analyzed with these variables. For instance, TERTp mutations were more prevalent in patients who did not receive chemotherapy. Consequently, the predictive effect previously attributed to TERTp status could actually be attributed to the impact of chemotherapy, indicating a potential confounding effect.

#### ERN1 mutations

Based on recent studies, we performed a targeted sequencing of an additional gene that we are currently studying: ERN1, encoding for the protein IRE1α (Lhomond et al., 2018). We sequenced the regions involving all the brain cancer-related mutations in the ERN1 gene known to date (Barthel et al., 2019; Vaubel et al., 2020; L. B. Wang et al., 2021; Weinstein et al., 2013); however, we did not identify any of the previously reported somatic mutations. Nonetheless, we found a likely germinal variant of ERN1 (g.chr17:64044182G>T, rs139229826; mean AF: 52% (18-99)) in 8 out of 70 patients (11%), that despite being a single-nucleotide polymorphism (SNP) previously reported in a healthy population (*ALFA: Allele Frequency Aggregator*, n.d.; Manolio, 2009; Matise et al., 2011), has a massive ethnic disparity between healthy Latin-Americans and the rest of the world (*ALFA: Allele Frequency Aggregator*, n.d.). Although this variant did not show a prognostic value in our cohort, it had not been previously described in glioblastoma patients and its exact impact on IRE1α function and glioblastoma is currently unknown.

IRE1α is among the kinases most mutated in cancer (Greenman et al., 2007). IRE1a signaling has been associated with different mechanisms involved in glioblastoma progression, including tumor growth, angiogenesis, invasions (Auf et al., 2010; Drogat et al., 2007; Jabouille et al., 2015; Le Reste et al., 2020a; Lhomond et al., 2018; Obacz et al., 2017) and several ERN1 mutations have been described in the context of glioblastoma (Barthel et al., 2019; Vaubel et al., 2020; L. B. Wang et al., 2021; Weinstein et al., 2013). IRE1α is an endoplasmic reticulum (ER) stress sensor that plays a crucial role in the unfolded protein response (UPR), regulating cell survival and adaptation during stress through the unconventional splicing of X-box binding protein 1 (XBP1) mRNA ref. IRE1α plays a crucial role in cancer by promoting cell survival under stress conditions and in response to oncogene inhibition by maintaining protein homeostasis (proteostasis) (Lv et al., 2023a; Obacz et al., 2017). This enables tumor growth and resistance to therapies, highlighting IRE1α as a potential target for cancer treatment, as supported by several preclinical studies clinical trials using MKC8866, a potent and specific IRE1α inhibitor (Le Reste et al., 2020b; Lv et al., 2023b; Pelizzari-Raymundo et al., 2023; Sheng et al., 2019).

#### Insights into ERN1 L914M mutation by 3D modelling structural changes

The ERN1 variant rs139229826 causes the L914M (lysine for methionine) substitution in the IRE1α protein. To define putative effects of this substitution in the function of IRE1α, we generated in silico 3D models using the human IRE1α cytosolic region as template (PDB 3P23). The L914 residue is highly conserved across various species (Supplementary Fig. 1). This mutation is particularly intriguing because the L914 residue is in close proximity to the essential catalytic residue K907, which is crucial for IRE1α’s RNase activity (Fig. 3A-B) (Ali et al., 2011). The substitution of L with M maintains the hydrophobic nature of the side chain, suggesting that it’s likely to be structurally well-tolerated by the protein. Given its proximity to the catalytic K907 residue, crucial for RNase activity, we performed a molecular dynamics simulation comparing IRE1α wildtype and IRE1α L914M (Fig 3C and D). Our findings suggest that the L914M mutation produces a conformational change in the K907 as a result of the disruption of an ionic bond between K907 and D885, suggesting that this substitution will have a notable impact on RNase activity. Additionally, this change in the amino acid composition could potentially alter the charge of the residue, leading to the incorporation of an amino acid susceptible to oxidation (Kim et al., 2014). Methionine contains a sulfur atom that can readily undergo oxidation, forming the corresponding sulphoxide (MetO), transforming a nonpolar side chain into a highly polar one. Altogether, the L914M mutation in IRE1α is of interest due to its proximity to the catalytic K907 residue, however, the functional significance to glioblastoma remains to be confirmed in cell culture models of GBM.

## Discussion

Numerous studies have elucidated variations in molecular markers associated with various cancer types in Latin American populations (Gonzalez-Pons & Cruz-Correa, 2019; Patel et al., 2019; Raez et al., 2020; Zevallos et al., 2020). However, Hispanic and Latino populations have been largely underrepresented in glioblastoma research (Guerrero et al., 2018). While some studies have indicated disparities in both the incidence and prognosis of glioblastoma among individuals of Latino and non-Latino backgrounds (Patel et al., 2019; Walsh et al., 2023), no comprehensive comparisons have been made regarding the molecular features of gliomas between Latin American and other populations. Notably, two previous single-center studies have specifically examined the prevalence of IDH1 mutations and MGMT methylation among Caucasian and Hispanic/Latino populations in the United States, which showed no racial influence in their mutation frequency (Shabihkhani et al., 2017; T. Wang et al., 2014).

Public health measures aimed at the implementation of diagnostic and therapeutic protocols mostly rely on international databases and clinical trials. Furthermore, recognizing the molecular features of different ethnic groups also allows for a better design of upcoming clinical trials. Here we conducted the first comprehensive molecular profiling of a Latin American glioblastoma cohort to establish a foundation for potential international comparisons. Our study on high-grade gliomas in a Chilean population reveals comparable findings with other cohorts from North American, European and Asian origin, suggesting a universal relevance of key molecular markers such as IDH, TERTp, TP53 and PTEN mutations as well as EGFR amplification and CDKN2A deletions (Brennan et al., 2013; Ceccarelli et al., 2016; Verhaak et al., 2010). Novel TP53 and PTEN variants were identified, determined to be likely pathogenic using in silico predictions, and further supported by the presence of similar alterations in other types of cancer, particularly in the case of frameshift mutations (Sherry et al., 1999). Mutations in the ATRX and SMARCAL1 genes related to Alternative Lengthening of Telomeres (ALT) were observed less frequently compared to findings in other studies (Diplas et al., 2018; Haase et al., 2018; Nandakumar et al., 2017). Nevertheless, the disparity in our results might be attributed, at least partially, to the design of our panel and its partial coverage of some of the most commonly mutated regions within these genes. In addition, lower-than-expected mutation rates were found in the NF1 gene.

These genetic insights underscore the importance of applying genomic techniques in diagnosis and expand the understanding of glioblastoma genetics, contributing to future classification and the development of possible treatment strategies. The prognostic value of these markers on patient survival was also evaluated, yielding results comparable to previous studies of non-Latin American and European populations (Killela et al., 2014; Nonoguchi et al., 2013; Yan et al., 2009).

Given the recent updates in the central nervous system tumor classification, the capacity to clinically identify mutations that were predominantly used in research settings in the past will become markedly important in the upcoming years. For instance, in our series, IDH sequencing allowed the re-categorization of a patient with negative IDH R132H IHC and a R172K mutation identified on NGS, moving from a glioblastoma to a grade IV IDH-mutant astrocytoma. Also, the relatively older age of patients with IDH mutations in this study suggests that the clinical use of DNA sequencing could be more relevant than currently considered in patients over 55 years old in our population. Similarly, our results re-categorized a patient with a *diffuse midline glioma, H3 K27-altered*, which is a pediatric-type tumor with a worse prognosis than glioblastoma (Louis et al., 2018). The current WHO classification allows for the first time to grade an astrocytic tumor based on molecular markers (TERTp mutations, EGFR amplification, chr7 gain/chr10 loss or CDKN2A deletion), which may inform specific therapeutic approaches regarding the use of adjuvant treatments such as radiotherapy and chemotherapy. While the inclusion of molecular data in our study did not lead to any modifications in tumor grading, it is worth noting that the primary beneficiary group for such information are individuals with lower grade gliomas, which were not the focus of our study.

Although the costs of sequencing have dramatically decreased over the last 15 years, the implementation and overall costs of the complete sequencing process remain very high to be used in most developing countries (Desai et al., 2021; Schwarze et al., 2019), making a simplified targeted sequencing strategy - like the one used in this study - an attractive alternative (Zacher et al., 2017). One of the main challenges in parallel mass sequencing is determining the sensitivity of the technique for low allelic frequency variants in tissues with low tumor cellularity. We optimized the conditions to reduce the risk of false negatives (DNA extraction from highly cellular tumor tissues, designing sequencing panels with high coverage depth, and setting low read number limits for known variants), but a standardized clinical implementation also requires the use of positive controls to establish the sensitivity and specificity of the sequencing strategy in a local context (Koboldt, 2020; Strom, 2016).

### The ERN variant rs139229826

ERN1 variants have been described in less than 2% of glioblastoma patients (Barthel et al., 2019; Vaubel et al., 2020; L. B. Wang et al., 2021; Weinstein et al., 2013). However, to the best of our knowledge, the rs139229826 variant present in 11% of our patients has not been previously reported in glioblastoma patients on any public database. Different population-based genomic studies including more than 400.000 subjects have found a global allelic frequency for the rs139229826 variant of 1% in healthy populations (Auton et al., 2015; Chen et al., 2022; Taliun et al., 2021). Yet, ethnic-specific analyses show a wide range of allelic frequencies going from 1 every 10.000 cases in European populations (*ALFA: Allele Frequency Aggregator*, n.d.; Auton et al., 2015), up to 9.1% (Manolio, 2009; Matise et al., 2011) for a group of participants of South American descent (n = 991; BioSample: SAMN10868969), and 9.7% (*ALFA: Allele Frequency Aggregator*, n.d.) for a different group of Latin American individuals with mostly European and Native American Ancestry (n = 632; BioSample: SAMN10492700), accounting for almost a 1000-fold difference. The rs139229826 variant of the ERN1 gene results in a mutant IRE1α protein with a L914M aminoacidic substitution. Our modeling of mutated IRE1α indicated that L914 residue is spatially close to K907, a key catalytic residue, which may affect the XBP-1 mRNA reaction as discussed.

Germline mutations are well-recognized for their role in cancer predisposition (Chatrath et al., 2020) and prediction of adverse events of certain chemotherapies (pharmacogenetics) (Kaehler & Cascorbi, 2019; O’Donnell & Ratain, 2012). In contrast, evidence regarding the predictive value of germline mutations in drug efficacy or their possible effects in the pathophysiology of tumor progression is widely lacking (Vali-Pour et al., 2022). Intriguingly, due to the nature of the protein change induced by rs139229826, it is likely that the L914M substitution is well tolerated under normal cellular conditions, but it might affect IRE1 regulatory functions under cellular stress. Importantly, the activity of IRE1 has been shown to be relevant for glioblastoma progression in mouse models and a relevant drug target (Auf et al., 2010; Drogat et al., 2007; Jabouille et al., 2015; Lhomond et al., 2018), correlating with poor glioblastoma patient prognosis (Lhomond et al., 2018; Pluquet et al., 2013).

### Study limitations

We would like to acknowledge some limitations of the current study: (i) Heterogeneous access to chemotherapy and radiotherapy generates a heterogeneous population to associate genetic mutations with patient survival. Although it allows us to retrospectively appreciate the possible effects of these treatments on clinical outcomes, it might mask the contributions of some additional prognostic factors in multivariate analyses (i.e. TERTp mutations) and prevents the comparison of clinical outcomes between this cohort and other international cohorts. The limited access to adjuvant therapies was a frequent occurrence in Latin-American health systems during the last decade. (ii) The extent of resection was not systematically measured with immediate postoperative magnetic resonance imaging. As evidence of the impact of small residual tumor on clinical outcomes was controversial at the time, it was deemed reasonable to estimate the resection based on postoperative computed tomographies (CT) and intraoperative surgeon’s judgement. (iii) O(6)-methylguanine-DNA methyltransferase (MGMT) promoter methylation was not determined because it is not currently included in the WHO classification system. However, several studies support its predictive value for temozolomide response (Binabaj et al., 2018; Brandner et al., 2021), and it might round- up the molecular study of future cohorts. (iv) The determination of gene copy number changes using amplicon sequencing as opposed to whole-genome sequencing has inherent limitations, mainly associated with coverage heterogeneity in different regions of interest (Petrackova et al., 2019). These limitations were mitigated with bioinformatic processing of the data (see methods), yet a formal comparison with a gold standard technique was not performed because this study was designed for research purposes.

## Conclusion

Our high-grade glioma cohort from Latin America exhibited a comparable incidence of IDH, TERTp, PTEN, and TP53 mutations to those reported in other studies with different ethnic origin. Conversely, NF1, ATRX and SMARCAL1 mutations demonstrated a lower frequency than expected. It’s important to note that some ATRX and SMARCAL1 mutation sites were not covered by our sequencing design. The correlation between IDH and TERTp mutations with patient survival indicated a similar association as reported in previous studies in populations of non-Latino American, European and Asian origin. Discovery of the ethnic-specific ERN1 variant rs139229826, not previously reported in glioblastoma, offers new prospects for future research. Our study contributes to the advancement of future international comparative studies and potential strategies for treating glioblastoma. Our findings illuminate the multifaceted nature of glioblastoma, underscoring its complex molecular origins and the distinctive genetic diversities that exist among various populations. We emphasize the importance of ensuring equitable access to advanced molecular diagnostics, shedding light on the need and approaches for the widespread adoption of state-of-the-art technologies in healthcare systems.

## Data Availability

All data produced in the present study are available upon reasonable request to the authors

## Acknowledgments

We thank Claudia Sepulveda and Susana Manriquez for lab managing and administrative accounting. We also thank Pamela Moller for her technical support regarding immunohistochemistry and sample management. This work was primarily funded by ANID/FONDECYT 1220573 (CH), FONDAP program 15150012 (CH), and funds from the Metropolitan East Health Service and the Department of Neurological Sciences of the University of Chile. We also thank support by the U.S. Air Force Office of Scientific Research FA9550-21-1-0096, Department of Defense grant W81XWH2110960, Millennium Institute P09-015-F (CH), ANID/FONDEF ID1ID22I10120, ANID/FONDECYT 11180825 (HH), postdoctoral ANID/FONDECYT 3210294 (PP), doctoral ANID/FONDECYT fellowship (RF), ANID/NAM22I0057 and Swiss Consolidation Grant -The Leading House for the Latin American Region (CH). This study is dedicated to Virgina Flores Kehr (CH family member) who died of glioblastoma during the course of the study.

## Author Contributions

Project conceptualization: RF, JM, and CH;

Methodology: RF, JM and CH;

Formal analysis: RF, MS, GC, PP, MA and SR;

Investigation: RF, HU, PP, BF, CS, DR, RM and CT;

Software: MS;

Data curation: RF, HU, PP;

Resources: CS and CH;

Writing – Original draft: RF, HU and PP;

Writing – Review and editing: all authors;

Supervision and project administration: JM and CH;

Funding acquisition: RF, DR, JM and CH.

## Supplementary material

**Supplementary Figure 1. Protein sequence alignment of IRE1**α **orthologs**. In the upper panel, variations in residues between species are depicted by comparing each residue at a specific position to the column consensus within the N-terminal region of IRE1α (residues 832 to 936 of the human IRE1α). The intensity of red shading reflects the extent of difference from residues in other rows at that position, with darker shades indicating greater divergence. The lower panel showcases differences relative to the human IRE1α protein sequence. We highlight the conservation of the critical leucine (L) at position 914 across diverse species.

**Supplementary Table 1. Target regions of the panel design.** The target coordinates for Ampliseq™ and Truseq™ sequencing strategies are listed. The Ampliseq™ design included 339 amplicons with an average length of 134 base pairs (ranging from 125 to 140 base pairs) and covered 15,596 base pairs, including the following genes: IDH1, IDH2, H3F3A, H3C2, EGFR, CDKN2A, ATRX, SMARCAL1, BRAF, TP53, PTEN, and ERN1. The Truseq™ design included 25 amplicons with an average length of 230 base pairs (ranging from 192 to 266 base pairs) that covered 5,750 base pairs, including the following genes: H3F3A, IDH1, IDH2, SMARCAL1, CDKN2A, ERN1.

**Supplementary Table 2. Summary of genomic variants.** Variant coordinates, classification, allelic frequencies, protein changes and samples presenting with each variant are provided. 61 unique variants were identified in 66 out of 70 patients.

**Supplementary Table 3. Summary of genomic variants coverage.** The sequencing depths (number of sequencing reads) over each variant are shown for all samples. Average and median coverage for each variant are also provided.

