## Supplementary material for "Characterization of the mutational landscape of high-grade gliomas in a Latin American cohort"

Supplementary Figure 1

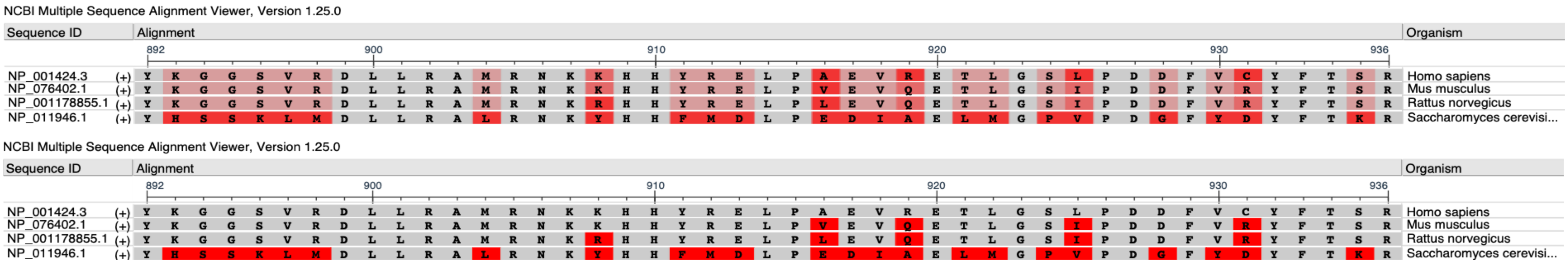

Supplementary Table 11

| Ampliseq primers | | | | |
|---|---|---|---|---|
| AmpliconId | Chromosome | Gene | Start_Coordinate | End_Coordinate |
| AMPL656428 | chr1 | H3F3A | 226064194 | 226064278 |
| AMPL656470 | chr1 | H3F3A | 226064481 | 226064549 |
| AMPL656451 | chr2 | IDH1 | 208248213 | 208248285 |
| AMPL656452 | chr2 | IDH1 | 208248274 | 208248341 |
| AMPL656453 | chr2 | IDH1 | 208248326 | 208248406 |
| AMPL368995 | chr2 | IDH1 | 208248397 | 208248485 |
| AMPL368171 | chr2 | IDH1 | 208248484 | 208248563 |
| AMPL368172 | chr2 | IDH1 | 208248552 | 208248631 |
| AMPL368173 | chr2 | IDH1 | 208248603 | 208248677 |
| AMPL380375 | chr5 | TERT | 1294941 | 1295036 |
| AMPL499877 | chr6 | HIST1H3B | 26031625 | 26031713 |
| AMPL499878 | chr6 | HIST1H3B | 26031704 | 26031795 |
| AMPL499879 | chr6 | HIST1H3B | 26031773 | 26031863 |
| AMPL499880 | chr6 | HIST1H3B | 26031843 | 26031934 |
| AMPL499881 | chr6 | HIST1H3B | 26031923 | 26032022 |
| AMPL499882 | chr6 | HIST1H3B | 26031954 | 26032047 |
| AMPL499883 | chr6 | HIST1H3B | 26032047 | 26032122 |
| AMPL658497 | chr7 | EGFR | 55019238 | 55019338 |
| AMPL30339 | chr7 | EGFR | 55019255 | 55019353 |
| AMPL30340 | chr7 | EGFR | 55019380 | 55019473 |
| AMPL865449 | chr7 | EGFR | 55019383 | 55019487 |
| AMPL656522 | chr7 | EGFR | 55142185 | 55142259 |
| AMPL656523 | chr7 | EGFR | 55142237 | 55142317 |
| AMPL30327 | chr7 | EGFR | 55142308 | 55142391 |
| AMPL30328 | chr7 | EGFR | 55142383 | 55142466 |
| AMPL656532 | chr7 | EGFR | 55142455 | 55142523 |
| AMPL865444 | chr7 | EGFR | 55143248 | 55143344 |
| AMPL656508 | chr7 | EGFR | 55143337 | 55143413 |
| AMPL656509 | chr7 | EGFR | 55143402 | 55143476 |
| AMPL656510 | chr7 | EGFR | 55143465 | 55143555 |
| AMPL30329 | chr7 | EGFR | 55146535 | 55146627 |
| AMPL31746 | chr7 | EGFR | 55146616 | 55146712 |
| AMPL658500 | chr7 | EGFR | 55146701 | 55146784 |
| AMPL656496 | chr7 | EGFR | 55146773 | 55146844 |
| AMPL656542 | chr7 | EGFR | 55151182 | 55151267 |
| AMPL656543 | chr7 | EGFR | 55151256 | 55151344 |
| AMPL30293 | chr7 | EGFR | 55151323 | 55151402 |
| AMPL865441 | chr7 | EGFR | 55152458 | 55152551 |
| AMPL865442 | chr7 | EGFR | 55152551 | 55152647 |
| AMPL31744 | chr7 | EGFR | 55152632 | 55152718 |
| AMPL658503 | chr7 | EGFR | 55152673 | 55152751 |
| AMPL30302 | chr7 | EGFR | 55153932 | 55154020 |
| AMPL30303 | chr7 | EGFR | 55154009 | 55154106 |

| | | | | |
|---|---|---|---|---|
| AMPL30304 | chr7 | EGFR | 55154078 | 55154171 |
| AMPL30305 | chr7 | EGFR | 55154127 | 55154221 |
| AMPL656497 | chr7 | EGFR | 55155624 | 55155717 |
| AMPL656498 | chr7 | EGFR | 55155708 | 55155796 |
| AMPL30296 | chr7 | EGFR | 55155738 | 55155828 |
| AMPL30297 | chr7 | EGFR | 55155817 | 55155915 |
| AMPL30298 | chr7 | EGFR | 55155904 | 55156000 |
| AMPL656539 | chr7 | EGFR | 55156406 | 55156494 |
| AMPL540285 | chr7 | EGFR | 55156483 | 55156558 |
| AMPL656540 | chr7 | EGFR | 55156527 | 55156602 |
| AMPL865437 | chr7 | EGFR | 55156591 | 55156664 |
| AMPL865438 | chr7 | EGFR | 55156624 | 55156711 |
| AMPL865439 | chr7 | EGFR | 55156697 | 55156782 |
| AMPL865440 | chr7 | EGFR | 55156774 | 55156859 |
| AMPL656518 | chr7 | EGFR | 55156848 | 55156927 |
| AMPL30261 | chr7 | EGFR | 55157590 | 55157662 |
| AMPL30262 | chr7 | EGFR | 55157651 | 55157742 |
| AMPL30263 | chr7 | EGFR | 55157731 | 55157803 |
| AMPL30277 | chr7 | EGFR | 55160054 | 55160143 |
| AMPL30278 | chr7 | EGFR | 55160132 | 55160207 |
| AMPL30279 | chr7 | EGFR | 55160194 | 55160272 |
| AMPL30280 | chr7 | EGFR | 55160240 | 55160327 |
| AMPL30281 | chr7 | EGFR | 55160317 | 55160400 |
| AMPL31736 | chr7 | EGFR | 55161412 | 55161509 |
| AMPL31737 | chr7 | EGFR | 55161498 | 55161599 |
| AMPL31738 | chr7 | EGFR | 55161588 | 55161665 |
| AMPL656476 | chr7 | EGFR | 55161628 | 55161713 |
| AMPL30275 | chr7 | EGFR | 55163679 | 55163765 |
| AMPL47950 | chr7 | EGFR | 55163751 | 55163834 |
| AMPL656483 | chr7 | EGFR | 55163774 | 55163868 |
| AMPL30334 | chr7 | EGFR | 55165208 | 55165283 |
| AMPL30335 | chr7 | EGFR | 55165272 | 55165350 |
| AMPL30336 | chr7 | EGFR | 55165316 | 55165400 |
| AMPL656502 | chr7 | EGFR | 55165387 | 55165463 |
| AMPL656503 | chr7 | EGFR | 55165453 | 55165537 |
| AMPL656484 | chr7 | EGFR | 55168356 | 55168424 |
| AMPL656485 | chr7 | EGFR | 55168410 | 55168475 |
| AMPL656486 | chr7 | EGFR | 55168441 | 55168519 |
| AMPL30246 | chr7 | EGFR | 55168508 | 55168569 |
| AMPL656515 | chr7 | EGFR | 55170142 | 55170226 |
| AMPL656516 | chr7 | EGFR | 55170206 | 55170294 |
| AMPL30265 | chr7 | EGFR | 55170283 | 55170369 |
| AMPL30266 | chr7 | EGFR | 55170358 | 55170446 |
| AMPL30267 | chr7 | EGFR | 55170444 | 55170540 |
| AMPL406048 | chr7 | EGFR | 55170529 | 55170626 |

| | | | | |
|---|---|---|---|---|
| AMPL406049 | chr7 | EGFR | 55170626 | 55170720 |
| AMPL406054 | chr7 | EGFR | 55171037 | 55171108 |
| AMPL656491 | chr7 | EGFR | 55171046 | 55171118 |
| AMPL30247 | chr7 | EGFR | 55171142 | 55171222 |
| AMPL656489 | chr7 | EGFR | 55171211 | 55171304 |
| AMPL30299 | chr7 | EGFR | 55172894 | 55172986 |
| AMPL31745 | chr7 | EGFR | 55172983 | 55173083 |
| AMPL30301 | chr7 | EGFR | 55173097 | 55173194 |
| AMPL30269 | chr7 | EGFR | 55173869 | 55173961 |
| AMPL30270 | chr7 | EGFR | 55173950 | 55174031 |
| AMPL30271 | chr7 | EGFR | 55174018 | 55174106 |
| AMPL656519 | chr7 | EGFR | 55174060 | 55174143 |
| AMPL656536 | chr7 | EGFR | 55174592 | 55174673 |
| AMPL656537 | chr7 | EGFR | 55174659 | 55174735 |
| AMPL656538 | chr7 | EGFR | 55174724 | 55174800 |
| AMPL865433 | chr7 | EGFR | 55174789 | 55174871 |
| AMPL656488 | chr7 | EGFR | 55174866 | 55174946 |
| AMPL31739 | chr7 | EGFR | 55181203 | 55181293 |
| AMPL31740 | chr7 | EGFR | 55181281 | 55181371 |
| AMPL31741 | chr7 | EGFR | 55181365 | 55181451 |
| AMPL658498 | chr7 | EGFR | 55181440 | 55181522 |
| AMPL656471 | chr7 | EGFR | 55191589 | 55191679 |
| AMPL30322 | chr7 | EGFR | 55191675 | 55191769 |
| AMPL30323 | chr7 | EGFR | 55191715 | 55191799 |
| AMPL30324 | chr7 | EGFR | 55191788 | 55191869 |
| AMPL30325 | chr7 | EGFR | 55191858 | 55191935 |
| AMPL30289 | chr7 | EGFR | 55192713 | 55192792 |
| AMPL30290 | chr7 | EGFR | 55192778 | 55192854 |
| AMPL30291 | chr7 | EGFR | 55192833 | 55192915 |
| AMPL656533 | chr7 | EGFR | 55198570 | 55198649 |
| AMPL30318 | chr7 | EGFR | 55198638 | 55198724 |
| AMPL30319 | chr7 | EGFR | 55198702 | 55198779 |
| AMPL30320 | chr7 | EGFR | 55198768 | 55198856 |
| AMPL30321 | chr7 | EGFR | 55198845 | 55198936 |
| AMPL656480 | chr7 | EGFR | 55200178 | 55200267 |
| AMPL47948 | chr7 | EGFR | 55200256 | 55200347 |
| AMPL865432 | chr7 | EGFR | 55200337 | 55200432 |
| AMPL30274 | chr7 | EGFR | 55200425 | 55200513 |
| AMPL656493 | chr7 | EGFR | 55201089 | 55201156 |
| AMPL656494 | chr7 | EGFR | 55201145 | 55201225 |
| AMPL656495 | chr7 | EGFR | 55201187 | 55201271 |
| AMPL47943 | chr7 | EGFR | 55201260 | 55201349 |
| AMPL47944 | chr7 | EGFR | 55201294 | 55201375 |
| AMPL656487 | chr7 | EGFR | 55201364 | 55201434 |
| AMPL656528 | chr7 | EGFR | 55201654 | 55201732 |

| | | | | |
|---|---|---|---|---|
| AMPL656529 | chr7 | EGFR | 55201713 | 55201786 |
| AMPL30256 | chr7 | EGFR | 55201774 | 55201845 |
| AMPL656513 | chr7 | EGFR | 55201808 | 55201879 |
| AMPL656514 | chr7 | EGFR | 55201879 | 55201949 |
| AMPL30252 | chr7 | EGFR | 55202452 | 55202545 |
| AMPL30253 | chr7 | EGFR | 55202535 | 55202633 |
| AMPL30254 | chr7 | EGFR | 55202559 | 55202657 |
| AMPL656506 | chr7 | EGFR | 55205158 | 55205240 |
| AMPL656507 | chr7 | EGFR | 55205229 | 55205312 |
| AMPL30307 | chr7 | EGFR | 55205312 | 55205394 |
| AMPL30308 | chr7 | EGFR | 55205331 | 55205420 |
| AMPL30309 | chr7 | EGFR | 55205412 | 55205493 |
| AMPL30310 | chr7 | EGFR | 55205437 | 55205526 |
| AMPL30311 | chr7 | EGFR | 55205515 | 55205590 |
| AMPL30312 | chr7 | EGFR | 55205575 | 55205653 |
| AMPL656455 | chr7 | BRAF | 140753110 | 140753187 |
| AMPL865434 | chr7 | BRAF | 140753200 | 140753280 |
| AMPL402786 | chr7 | BRAF | 140753275 | 140753352 |
| AMPL385965 | chr7 | BRAF | 140753341 | 140753415 |
| AMPL473617 | chr7 | BRAF | 140753377 | 140753443 |
| AMPL473618 | chr7 | BRAF | 140753437 | 140753509 |
| AMPL656436 | chr7 | BRAF | 140753477 | 140753549 |
| AMPL656437 | chr7 | BRAF | 140753542 | 140753609 |
| AMPL656441 | chr7 | BRAF | 140781560 | 140781643 |
| AMPL656442 | chr7 | BRAF | 140781595 | 140781676 |
| AMPL30217 | chr7 | BRAF | 140781668 | 140781736 |
| AMPL399513 | chr9 | CDKN2A | 21968113 | 21968200 |
| AMPL865435 | chr9 | CDKN2A | 21968189 | 21968274 |
| AMPL865436 | chr9 | CDKN2A | 21968263 | 21968337 |
| AMPL865450 | chr9 | CDKN2A | 21968648 | 21968746 |
| AMPL362492 | chr9 | CDKN2A | 21968743 | 21968826 |
| AMPL398554 | chr9 | CDKN2A | 21970840 | 21970931 |
| AMPL865446 | chr9 | CDKN2A | 21970896 | 21970977 |
| AMPL865447 | chr9 | CDKN2A | 21971236 | 21971328 |
| AMPL48429 | chr9 | CDKN2A | 21974428 | 21974512 |
| AMPL48430 | chr9 | CDKN2A | 21974492 | 21974574 |
| AMPL48431 | chr9 | CDKN2A | 21974533 | 21974616 |
| AMPL48432 | chr9 | CDKN2A | 21974605 | 21974695 |
| AMPL48433 | chr9 | CDKN2A | 21974684 | 21974785 |
| AMPL865431 | chr9 | CDKN2A | 21974938 | 21975026 |
| AMPL48437 | chr9 | CDKN2A | 21994069 | 21994166 |
| AMPL48438 | chr9 | CDKN2A | 21994155 | 21994248 |
| AMPL48439 | chr9 | CDKN2A | 21994199 | 21994296 |
| AMPL48440 | chr9 | CDKN2A | 21994280 | 21994375 |
| AMPL865443 | chr9 | CDKN2A | 21994319 | 21994412 |

| | | | | |
|---|---|---|---|---|
| AMPL30705 | chr10 | PTEN | 87864406 | 87864493 |
| AMPL48109 | chr10 | PTEN | 87864483 | 87864564 |
| AMPL656432 | chr10 | PTEN | 87864553 | 87864625 |
| AMPL593000 | chr10 | PTEN | 87893903 | 87893979 |
| AMPL30723 | chr10 | PTEN | 87894007 | 87894081 |
| AMPL30724 | chr10 | PTEN | 87894076 | 87894158 |
| AMPL656427 | chr10 | PTEN | 87894130 | 87894202 |
| AMPL592997 | chr10 | PTEN | 87925429 | 87925496 |
| AMPL30716 | chr10 | PTEN | 87925485 | 87925554 |
| AMPL30717 | chr10 | PTEN | 87925533 | 87925607 |
| AMPL656460 | chr10 | PTEN | 87925588 | 87925659 |
| AMPL656461 | chr10 | PTEN | 87930951 | 87931029 |
| AMPL656462 | chr10 | PTEN | 87930986 | 87931063 |
| AMPL30704 | chr10 | PTEN | 87931052 | 87931121 |
| AMPL592999 | chr10 | PTEN | 87931092 | 87931164 |
| AMPL656426 | chr10 | PTEN | 87931153 | 87931230 |
| AMPL656429 | chr10 | PTEN | 87932916 | 87932986 |
| AMPL865448 | chr10 | PTEN | 87932960 | 87933037 |
| AMPL30695 | chr10 | PTEN | 87933026 | 87933112 |
| AMPL30696 | chr10 | PTEN | 87933102 | 87933194 |
| AMPL30697 | chr10 | PTEN | 87933185 | 87933265 |
| AMPL30698 | chr10 | PTEN | 87933255 | 87933332 |
| AMPL656447 | chr10 | PTEN | 87951939 | 87952026 |
| AMPL656448 | chr10 | PTEN | 87952036 | 87952105 |
| AMPL656449 | chr10 | PTEN | 87952066 | 87952134 |
| AMPL30713 | chr10 | PTEN | 87952123 | 87952200 |
| AMPL30714 | chr10 | PTEN | 87952189 | 87952271 |
| AMPL30715 | chr10 | PTEN | 87952270 | 87952346 |
| AMPL592998 | chr10 | PTEN | 87957757 | 87957847 |
| AMPL48104 | chr10 | PTEN | 87957837 | 87957919 |
| AMPL48105 | chr10 | PTEN | 87957856 | 87957936 |
| AMPL30720 | chr10 | PTEN | 87957935 | 87958015 |
| AMPL30721 | chr10 | PTEN | 87957958 | 87958042 |
| AMPL30722 | chr10 | PTEN | 87958042 | 87958106 |
| AMPL656439 | chr10 | PTEN | 87960716 | 87960790 |
| AMPL656440 | chr10 | PTEN | 87960807 | 87960886 |
| AMPL30689 | chr10 | PTEN | 87960822 | 87960894 |
| AMPL30690 | chr10 | PTEN | 87960915 | 87960991 |
| AMPL30691 | chr10 | PTEN | 87960981 | 87961056 |
| AMPL30692 | chr10 | PTEN | 87961045 | 87961111 |
| AMPL656458 | chr10 | PTEN | 87965155 | 87965232 |
| AMPL30707 | chr10 | PTEN | 87965193 | 87965270 |
| AMPL30708 | chr10 | PTEN | 87965265 | 87965342 |
| AMPL30709 | chr10 | PTEN | 87965331 | 87965420 |
| AMPL30710 | chr10 | PTEN | 87965409 | 87965478 |

| | | | | |
|---|---|---|---|---|
| AMPL30711 | chr10 | PTEN | 87965467 | 87965542 |
| AMPL656423 | chr15 | IDH2 | 90088424 | 90088511 |
| AMPL373004 | chr15 | IDH2 | 90088509 | 90088609 |
| AMPL370993 | chr15 | IDH2 | 90088524 | 90088622 |
| AMPL370994 | chr15 | IDH2 | 90088619 | 90088702 |
| AMPL370997 | chr15 | IDH2 | 90088668 | 90088742 |
| AMPL141289 | chr17 | TP53 | 7669493 | 7669587 |
| AMPL141290 | chr17 | TP53 | 7669549 | 7669623 |
| AMPL141291 | chr17 | TP53 | 7669612 | 7669690 |
| AMPL141292 | chr17 | TP53 | 7669679 | 7669760 |
| AMPL405259 | chr17 | TP53 | 7670502 | 7670594 |
| AMPL46528 | chr17 | TP53 | 7670569 | 7670665 |
| AMPL46529 | chr17 | TP53 | 7670664 | 7670748 |
| AMPL46539 | chr17 | TP53 | 7673176 | 7673251 |
| AMPL656463 | chr17 | TP53 | 7673240 | 7673319 |
| AMPL656464 | chr17 | TP53 | 7673308 | 7673397 |
| AMPL405256 | chr17 | TP53 | 7673464 | 7673540 |
| AMPL405257 | chr17 | TP53 | 7673520 | 7673609 |
| AMPL405258 | chr17 | TP53 | 7673601 | 7673689 |
| AMPL405261 | chr17 | TP53 | 7673660 | 7673741 |
| AMPL46533 | chr17 | TP53 | 7673713 | 7673808 |
| AMPL405262 | chr17 | TP53 | 7673770 | 7673850 |
| AMPL405263 | chr17 | TP53 | 7673839 | 7673933 |
| AMPL656465 | chr17 | TP53 | 7673970 | 7674064 |
| AMPL865445 | chr17 | TP53 | 7674073 | 7674160 |
| AMPL46530 | chr17 | TP53 | 7674137 | 7674229 |
| AMPL368771 | chr17 | TP53 | 7674217 | 7674312 |
| AMPL405266 | chr17 | TP53 | 7674777 | 7674870 |
| AMPL46546 | chr17 | TP53 | 7674824 | 7674901 |
| AMPL140965 | chr17 | TP53 | 7674890 | 7674969 |
| AMPL140966 | chr17 | TP53 | 7674958 | 7675048 |
| AMPL46543 | chr17 | TP53 | 7674997 | 7675091 |
| AMPL60 | chr17 | TP53 | 7675068 | 7675160 |
| AMPL46544 | chr17 | TP53 | 7675128 | 7675207 |
| AMPL656469 | chr17 | TP53 | 7675182 | 7675268 |
| AMPL140961 | chr17 | TP53 | 7675902 | 7675997 |
| AMPL140962 | chr17 | TP53 | 7675975 | 7676067 |
| AMPL46536 | chr17 | TP53 | 7676067 | 7676165 |
| AMPL46537 | chr17 | TP53 | 7676090 | 7676185 |
| AMPL46538 | chr17 | TP53 | 7676184 | 7676277 |
| AMPL140964 | chr17 | TP53 | 7676267 | 7676363 |
| AMPL46548 | chr17 | TP53 | 7676300 | 7676390 |
| AMPL46549 | chr17 | TP53 | 7676397 | 7676492 |
| AMPL381025 | chr17 | TP53 | 7676464 | 7676563 |
| AMPL381026 | chr17 | TP53 | 7676552 | 7676642 |

| | | | | |
|---|---|---|---|---|
| AMPL866642 | chr17 | NF1 | 31156039 | 31156128 |
| AMPL47393 | chr17 | NF1 | 31169973 | 31170035 |
| AMPL47431 | chr17 | NF1 | 31181446 | 31181525 |
| AMPL866636 | chr17 | NF1 | 31182534 | 31182620 |
| AMPL47323 | chr17 | NF1 | 31206264 | 31206354 |
| AMPL866634 | chr17 | NF1 | 31206308 | 31206392 |
| AMPL368806 | chr17 | NF1 | 31226377 | 31226467 |
| AMPL47344 | chr17 | NF1 | 31226662 | 31226743 |
| AMPL866639 | chr17 | NF1 | 31227238 | 31227330 |
| AMPL47349 | chr17 | NF1 | 31229006 | 31229087 |
| AMPL47379 | chr17 | NF1 | 31229771 | 31229851 |
| AMPL76909 | chr17 | NF1 | 31230320 | 31230395 |
| AMPL866641 | chr17 | NF1 | 31232763 | 31232849 |
| AMPL866648 | chr17 | NF1 | 31235599 | 31235684 |
| AMPL866645 | chr17 | NF1 | 31261658 | 31261745 |
| AMPL866640 | chr17 | NF1 | 31327707 | 31327791 |
| AMPL866643 | chr17 | NF1 | 31327804 | 31327893 |
| AMPL866644 | chr17 | NF1 | 31334880 | 31334965 |
| AMPL47434 | chr17 | NF1 | 31334975 | 31335058 |
| AMPL866638 | chr17 | NF1 | 31338030 | 31338110 |
| AMPL47385 | chr17 | NF1 | 31338728 | 31338813 |
| AMPL866637 | chr17 | NF1 | 31340572 | 31340656 |
| AMPL866633 | chr17 | NF1 | 31350182 | 31350270 |
| AMPL866647 | chr17 | NF1 | 31356487 | 31356554 |
| AMPL593577 | chr17 | NF1 | 31357282 | 31357356 |
| AMPL866574 | chrX | ATRX | 77523207 | 77523290 |
| AMPL866564 | chrX | ATRX | 77523296 | 77523377 |
| AMPL866568 | chrX | ATRX | 77557541 | 77557616 |
| AMPL79778 | chrX | ATRX | 77558690 | 77558758 |
| AMPL372328 | chrX | ATRX | 77574281 | 77574352 |
| AMPL866550 | chrX | ATRX | 77593734 | 77593819 |
| AMPL866552 | chrX | ATRX | 77599423 | 77599510 |
| AMPL866549 | chrX | ATRX | 77599532 | 77599610 |
| AMPL866573 | chrX | ATRX | 77600466 | 77600541 |
| AMPL866556 | chrX | ATRX | 77616615 | 77616684 |
| AMPL372321 | chrX | ATRX | 77618827 | 77618913 |
| AMPL866535 | chrX | ATRX | 77620419 | 77620491 |
| AMPL866533 | chrX | ATRX | 77633193 | 77633267 |
| AMPL866529 | chrX | ATRX | 77634595 | 77634679 |
| AMPL866546 | chrX | ATRX | 77635973 | 77636052 |
| AMPL79791 | chrX | ATRX | 77652092 | 77652170 |
| AMPL866570 | chrX | ATRX | 77654099 | 77654178 |
| AMPL866532 | chrX | ATRX | 77656524 | 77656609 |
| AMPL866554 | chrX | ATRX | 77656620 | 77656701 |
| AMPL372325 | chrX | ATRX | 77663426 | 77663508 |

| | | | | |
|---|---|---|---|---|
| AMPL866548 | chrX | ATRX | 77663505 | 77663589 |
| AMPL866566 | chrX | ATRX | 77664666 | 77664749 |
| AMPL866547 | chrX | ATRX | 77681702 | 77681789 |
| AMPL866538 | chrX | ATRX | 77681896 | 77681967 |
| AMPL866562 | chrX | ATRX | 77682075 | 77682160 |
| AMPL866553 | chrX | ATRX | 77682241 | 77682328 |
| AMPL866542 | chrX | ATRX | 77682385 | 77682475 |
| AMPL866559 | chrX | ATRX | 77682488 | 77682580 |
| AMPL866537 | chrX | ATRX | 77682557 | 77682640 |
| AMPL866563 | chrX | ATRX | 77682713 | 77682798 |
| AMPL866571 | chrX | ATRX | 77682785 | 77682869 |
| AMPL866557 | chrX | ATRX | 77682873 | 77682949 |
| AMPL866544 | chrX | ATRX | 77683025 | 77683092 |
| AMPL866551 | chrX | ATRX | 77683222 | 77683295 |
| AMPL372258 | chrX | ATRX | 77683563 | 77683633 |
| AMPL866531 | chrX | ATRX | 77683606 | 77683692 |
| AMPL866545 | chrX | ATRX | 77683797 | 77683880 |
| AMPL866543 | chrX | ATRX | 77683900 | 77683966 |
| AMPL866561 | chrX | ATRX | 77684063 | 77684152 |
| AMPL372267 | chrX | ATRX | 77684133 | 77684220 |
| AMPL866555 | chrX | ATRX | 77684263 | 77684347 |
| AMPL866540 | chrX | ATRX | 77684326 | 77684414 |
| AMPL866575 | chrX | ATRX | 77684424 | 77684511 |
| AMPL866567 | chrX | ATRX | 77684537 | 77684622 |
| AMPL372352 | chrX | ATRX | 77684913 | 77684999 |
| **Truseq amplicons** | | | | |
| AmpliconId | Chromosome | Gene | Start_Coordinate | End_Coordinate |
| hH3F3A | chr1 | H3F3A | 226064379 | 226064572 |
| hIDH1 | chr2 | IDH1 | 208248354 | 208248575 |
| hSMARCAL1-645 | chr2 | SMARCAL1 | 216450877 | 216451079 |
| hSMARCAL1-793 | chr2 | SMARCAL1 | 216475279 | 216475481 |
| hSMARCAL1-_945 | chr2 | SMARCAL1 | 216482872 | 216483106 |
| hTERT_Set_2 | chr5 | TERT | 1295037 | 1295278 |
| hCDKN2A_Ex_3A | chr9 | CDKN2A | 21967691 | 21967941 |
| hCDKN2A_Ex_3B | chr9 | CDKN2A | 21967904 | 21968143 |
| hCDKN2A_Ex_2A | chr9 | CDKN2A | 21970969 | 21971233 |
| hCDKN2A_Ex_1b | chr9 | CDKN2A | 21974961 | 21975163 |
| hIDH2 | chr15 | IDH2 | 90088556 | 90088800 |
| hIRE_p.P915R | chr17 | ERN1 | 64044131 | 64044366 |
| hIRE_R887C | chr17 | ERN1 | 64044859 | 64045070 |
| hIRE_V826M | chr17 | ERN1 | 64047849 | 64048076 |
| hIRE_Q780*/S769F | chr17 | ERN1 | 64048962 | 64049206 |
| hIRE_R627L | chr17 | ERN1 | 64054268 | 64054480 |
| hIRE_A414T | chr17 | ERN1 | 64057789 | 64058021 |
| hIRE_P380S | chr17 | ERN1 | 64060466 | 64060700 |

| | | | | |
|---|---|---|---|---|
| hIRE_p.P336L | chr17 | ERN1 | 64063890 | 64064124 |
| hIRE_p.V216= | chr17 | ERN1 | 64066713 | 64066930 |
| hIRE_X95_splice | chr17 | ERN1 | 64075190 | 64075429 |
| hIRE_X59_splice | chr17 | ERN1 | 64097972 | 64098193 |

### Supplementary Table 2

| Hugo_Symbol | Entrez_Gene_Id | Center | NCBI_Build | Chromosome | Start_Position | End_Position | Strand | Variant_Classification | Variant_Type | Reference_Allele | Tumor_Seq_Allele1 | Tumor_Seq_Allele2 | Tumor_Sample_Barcode | Protein_Change | Allelic_frequency | i_transcript_name |
|---|---|---|---|---|---|---|---|---|---|---|---|---|---|---|---|---|
| TERT | | Asenjo Neurosurgery Institute | hg38 | chr5 | 1295113 | 1295113 | + | 5'Flank | SNP | G | G | A | HGG_4 | | 0,089552239 | NM_198253.2 |
| TERT | | Asenjo Neurosurgery Institute | hg38 | chr5 | 1295113 | 1295113 | + | 5'Flank | SNP | G | G | A | HGG_6 | | 0,295774648 | NM_198253.2 |
| TERT | | Asenjo Neurosurgery Institute | hg38 | chr5 | 1295113 | 1295113 | + | 5'Flank | SNP | G | G | A | HGG_7 | | 0,453900709 | NM_198253.2 |
| TERT | | Asenjo Neurosurgery Institute | hg38 | chr5 | 1295113 | 1295113 | + | 5'Flank | SNP | G | G | A | HGG_10 | | 0,376425856 | NM_198253.2 |
| TERT | | Asenjo Neurosurgery Institute | hg38 | chr5 | 1295113 | 1295113 | + | 5'Flank | SNP | G | G | A | HGG_11 | | 0,3640553 | NM_198253.2 |
| TERT | | Asenjo Neurosurgery Institute | hg38 | chr5 | 1295113 | 1295113 | + | 5'Flank | SNP | G | G | A | HGG_12 | | 0,385714286 | NM_198253.2 |
| TERT | | Asenjo Neurosurgery Institute | hg38 | chr5 | 1295113 | 1295113 | + | 5'Flank | SNP | G | G | A | HGG_14 | | 0,396875 | NM_198253.2 |
| TERT | | Asenjo Neurosurgery Institute | hg38 | chr5 | 1295113 | 1295113 | + | 5'Flank | SNP | G | G | A | HGG_15 | | 0,38 | NM_198253.2 |
| TERT | | Asenjo Neurosurgery Institute | hg38 | chr5 | 1295113 | 1295113 | + | 5'Flank | SNP | G | G | A | HGG_16 | | 0,355191257 | NM_198253.2 |
| TERT | | Asenjo Neurosurgery Institute | hg38 | chr5 | 1295113 | 1295113 | + | 5'Flank | SNP | G | G | A | HGG_20 | | 0,464705882 | NM_198253.2 |
| TERT | | Asenjo Neurosurgery Institute | hg38 | chr5 | 1295113 | 1295113 | + | 5'Flank | SNP | G | G | A | HGG_21 | | 0,40397351 | NM_198253.2 |
| TERT | | Asenjo Neurosurgery Institute | hg38 | chr5 | 1295113 | 1295113 | + | 5'Flank | SNP | G | G | A | HGG_23 | | 0,26 | NM_198253.2 |
| TERT | | Asenjo Neurosurgery Institute | hg38 | chr5 | 1295113 | 1295113 | + | 5'Flank | SNP | G | G | A | HGG_24 | | 0,443298969 | NM_198253.2 |
| TERT | | Asenjo Neurosurgery Institute | hg38 | chr5 | 1295113 | 1295113 | + | 5'Flank | SNP | G | G | A | HGG_25 | | 0,362459547 | NM_198253.2 |
| TERT | | Asenjo Neurosurgery Institute | hg38 | chr5 | 1295113 | 1295113 | + | 5'Flank | SNP | G | G | A | HGG_26 | | 0,475274725 | NM_198253.2 |
| TERT | | Asenjo Neurosurgery Institute | hg38 | chr5 | 1295113 | 1295113 | + | 5'Flank | SNP | G | G | A | HGG_29 | | 0,238095238 | NM_198253.2 |
| TERT | | Asenjo Neurosurgery Institute | hg38 | chr5 | 1295113 | 1295113 | + | 5'Flank | SNP | G | G | A | HGG_30 | | 0,315315315 | NM_198253.2 |
| TERT | | Asenjo Neurosurgery Institute | hg38 | chr5 | 1295113 | 1295113 | + | 5'Flank | SNP | G | G | A | HGG_32 | | 0,44600939 | NM_198253.2 |
| TERT | | Asenjo Neurosurgery Institute | hg38 | chr5 | 1295113 | 1295113 | + | 5'Flank | SNP | G | G | A | HGG_35 | | 0,307692308 | NM_198253.2 |
| TERT | | Asenjo Neurosurgery Institute | hg38 | chr5 | 1295113 | 1295113 | + | 5'Flank | SNP | G | G | A | HGG_36 | | 0,25 | NM_198253.2 |
| TERT | | Asenjo Neurosurgery Institute | hg38 | chr5 | 1295113 | 1295113 | + | 5'Flank | SNP | G | G | A | HGG_40 | | 0,318918919 | NM_198253.2 |
| TERT | | Asenjo Neurosurgery Institute | hg38 | chr5 | 1295113 | 1295113 | + | 5'Flank | SNP | G | G | A | HGG_41 | | 0,251428571 | NM_198253.2 |
| TERT | | Asenjo Neurosurgery Institute | hg38 | chr5 | 1295113 | 1295113 | + | 5'Flank | SNP | G | G | A | HGG_43 | | 0,229357798 | NM_198253.2 |
| TERT | | Asenjo Neurosurgery Institute | hg38 | chr5 | 1295113 | 1295113 | + | 5'Flank | SNP | G | G | A | HGG_45 | | 0,368146214 | NM_198253.2 |
| TERT | | Asenjo Neurosurgery Institute | hg38 | chr5 | 1295113 | 1295113 | + | 5'Flank | SNP | G | G | A | HGG_48 | | 0,340909091 | NM_198253.2 |
| TERT | | Asenjo Neurosurgery Institute | hg38 | chr5 | 1295113 | 1295113 | + | 5'Flank | SNP | G | G | A | HGG_49 | | 0,365122616 | NM_198253.2 |
| TERT | | Asenjo Neurosurgery Institute | hg38 | chr5 | 1295113 | 1295113 | + | 5'Flank | SNP | G | G | A | HGG_51 | | 0,464454976 | NM_198253.2 |
| TERT | | Asenjo Neurosurgery Institute | hg38 | chr5 | 1295113 | 1295113 | + | 5'Flank | SNP | G | G | A | HGG_52 | | 0,49122807 | NM_198253.2 |
| TERT | | Asenjo Neurosurgery Institute | hg38 | chr5 | 1295113 | 1295113 | + | 5'Flank | SNP | G | G | A | HGG_53 | | 0,342105263 | NM_198253.2 |
| TERT | | Asenjo Neurosurgery Institute | hg38 | chr5 | 1295113 | 1295113 | + | 5'Flank | SNP | G | G | A | HGG_54 | | 0,079710145 | NM_198253.2 |
| TERT | | Asenjo Neurosurgery Institute | hg38 | chr5 | 1295113 | 1295113 | + | 5'Flank | SNP | G | G | A | HGG_58 | | 0,43062201 | NM_198253.2 |
| TERT | | Asenjo Neurosurgery Institute | hg38 | chr5 | 1295113 | 1295113 | + | 5'Flank | SNP | G | G | A | HGG_59 | | 0,326347305 | NM_198253.2 |
| TERT | | Asenjo Neurosurgery Institute | hg38 | chr5 | 1295113 | 1295113 | + | 5'Flank | SNP | G | G | A | HGG_60 | | 0,447941889 | NM_198253.2 |
| TERT | | Asenjo Neurosurgery Institute | hg38 | chr5 | 1295113 | 1295113 | + | 5'Flank | SNP | G | G | A | HGG_66 | | 0,157303371 | NM_198253.2 |
| TERT | | Asenjo Neurosurgery Institute | hg38 | chr5 | 1295113 | 1295113 | + | 5'Flank | SNP | G | G | A | HGG_67 | | 0,52688172 | NM_198253.2 |
| TERT | | Asenjo Neurosurgery Institute | hg38 | chr5 | 1295113 | 1295113 | + | 5'Flank | SNP | G | G | A | HGG_68 | | 0,458961474 | NM_198253.2 |
| TERT | | Asenjo Neurosurgery Institute | hg38 | chr5 | 1295113 | 1295113 | + | 5'Flank | SNP | G | G | A | HGG_69 | | 0,125944584 | NM_198253.2 |
| TERT | | Asenjo Neurosurgery Institute | hg38 | chr5 | 1295135 | 1295135 | + | 5'Flank | SNP | G | G | A | HGG_1 | | 0,357142857 | NM_198253.2 |
| TERT | | Asenjo Neurosurgery Institute | hg38 | chr5 | 1295135 | 1295135 | + | 5'Flank | SNP | G | G | A | HGG_18 | | 0,376106195 | NM_198253.2 |
| TERT | | Asenjo Neurosurgery Institute | hg38 | chr5 | 1295135 | 1295135 | + | 5'Flank | SNP | G | G | A | HGG_22 | | 0,241813602 | NM_198253.2 |
| TERT | | Asenjo Neurosurgery Institute | hg38 | chr5 | 1295135 | 1295135 | + | 5'Flank | SNP | G | G | A | HGG_39 | | 0,244318182 | NM_198253.2 |
| TERT | | Asenjo Neurosurgery Institute | hg38 | chr5 | 1295135 | 1295135 | + | 5'Flank | SNP | G | G | A | HGG_44 | | 0,542553191 | NM_198253.2 |
| TERT | | Asenjo Neurosurgery Institute | hg38 | chr5 | 1295135 | 1295135 | + | 5'Flank | SNP | G | G | A | HGG_47 | | 0,415147265 | NM_198253.2 |
| TERT | | Asenjo Neurosurgery Institute | hg38 | chr5 | 1295135 | 1295135 | + | 5'Flank | SNP | G | G | A | HGG_56 | | 0,105810398 | NM_198253.2 |
| TERT | | Asenjo Neurosurgery Institute | hg38 | chr5 | 1295135 | 1295135 | + | 5'Flank | SNP | G | G | A | HGG_62 | | 0,333333333 | NM_198253.2 |
| TERT | | Asenjo Neurosurgery Institute | hg38 | chr5 | 1295135 | 1295135 | + | 5'Flank | SNP | G | G | A | HGG_64 | | 0,261484099 | NM_198253.2 |
| TERT | | Asenjo Neurosurgery Institute | hg38 | chr5 | 1295135 | 1295135 | + | 5'Flank | SNP | G | G | A | HGG_70 | | 0,398981324 | NM_198253.2 |
| TP53 | 7157 | Asenjo Neurosurgery Institute | hg38 | chr17 | 7669615 | 7669615 | + | Frame_Shift_Del | DEL | T | T | - | HGG_49 | p.D393fs | 0,4503489 | NM_001276761.1 |
| TP53 | 7157 | Asenjo Neurosurgery Institute | hg38 | chr17 | 7673776 | 7673776 | + | Missense_Mutation | SNP | G | G | A | HGG_8 | p.R282W | 0,717871486 | NM_001276761.1 |
| TP53 | 7157 | Asenjo Neurosurgery Institute | hg38 | chr17 | 7673776 | 7673776 | + | Missense_Mutation | SNP | G | G | A | HGG_65 | p.R282W | 0,233176168 | NM_001276761.1 |
| TP53 | 7157 | Asenjo Neurosurgery Institute | hg38 | chr17 | 7673803 | 7673803 | + | Missense_Mutation | SNP | G | G | A | HGG_5 | p.R273C | 0,490497738 | NM_001276761.1 |
| TP53 | 7157 | Asenjo Neurosurgery Institute | hg38 | chr17 | 7673803 | 7673803 | + | Missense_Mutation | SNP | G | G | A | HGG_13 | p.R273C | 0,851030111 | NM_001276761.1 |
| TP53 | 7157 | Asenjo Neurosurgery Institute | hg38 | chr17 | 7673803 | 7673803 | + | Missense_Mutation | SNP | G | G | A | HGG_19 | p.R273C | 0,919324578 | NM_001276761.1 |
| TP53 | 7157 | Asenjo Neurosurgery Institute | hg38 | chr17 | 7673803 | 7673803 | + | Missense_Mutation | SNP | G | G | A | HGG_28 | p.R273C | 0,473471223 | NM_001276761.1 |
| TP53 | 7157 | Asenjo Neurosurgery Institute | hg38 | chr17 | 7673803 | 7673803 | + | Missense_Mutation | SNP | G | G | T | HGG_49 | p.R273S | 0,463312369 | NM_001276761.1 |
| TP53 | 7157 | Asenjo Neurosurgery Institute | hg38 | chr17 | 7673820 | 7673820 | + | Missense_Mutation | SNP | C | C | G | HGG_35 | p.R267P | 0,161111111 | NM_001276761.1 |
| TP53 | 7157 | Asenjo Neurosurgery Institute | hg38 | chr17 | 7673826 | 7673826 | + | Missense_Mutation | SNP | A | A | G | HGG_31 | p.L265P | 0,924686192 | NM_001276761.1 |
| TP53 | 7157 | Asenjo Neurosurgery Institute | hg38 | chr17 | 7674199 | 7674199 | + | Missense_Mutation | SNP | A | A | G | HGG_50 | p.I255T | 0,908256881 | NM_001276761.1 |
| TP53 | 7157 | Asenjo Neurosurgery Institute | hg38 | chr17 | 7674218 | 7674218 | + | Missense_Mutation | SNP | T | T | C | HGG_12 | p.R249G | 0,781818182 | NM_001276761.1 |
| TP53 | 7157 | Asenjo Neurosurgery Institute | hg38 | chr17 | 7674220 | 7674220 | + | Missense_Mutation | SNP | C | C | T | HGG_9 | p.R248Q | 0,373056995 | NM_001276761.1 |
| TP53 | 7157 | Asenjo Neurosurgery Institute | hg38 | chr17 | 7674220 | 7674220 | + | Missense_Mutation | SNP | C | C | T | HGG_43 | p.R248Q | 0,487616099 | NM_001276761.1 |
| TP53 | 7157 | Asenjo Neurosurgery Institute | hg38 | chr17 | 7674227 | 7674227 | + | Missense_Mutation | SNP | T | T | C | HGG_14 | p.M246V | 0,645348837 | NM_001276761.1 |
| TP53 | 7157 | Asenjo Neurosurgery Institute | hg38 | chr17 | 7674230 | 7674230 | + | Missense_Mutation | SNP | C | C | T | HGG_39 | p.G245S | 0,446376812 | NM_001276761.1 |
| TP53 | 7157 | Asenjo Neurosurgery Institute | hg38 | chr17 | 7674233 | 7674233 | + | Missense_Mutation | SNP | C | C | T | HGG_8 | p.G244S | 0,098360656 | NM_001276761.1 |
| TP53 | 7157 | Asenjo Neurosurgery Institute | hg38 | chr17 | 7674893 | 7674893 | + | Missense_Mutation | SNP | C | C | T | HGG_9 | p.R213Q | 0,400990099 | NM_001276761.1 |
| TP53 | 7157 | Asenjo Neurosurgery Institute | hg38 | chr17 | 7674894 | 7674894 | + | Nonsense_Mutation | SNP | G | G | A | HGG_41 | p.R213* | 0,361904762 | NM_001276761.1 |
| TP53 | 7157 | Asenjo Neurosurgery Institute | hg38 | chr17 | 7674904 | 7674905 | + | Frame_Shift_Del | DEL | TC | TC | - | HGG_27 | p.R209fs | 0,444487278 | NM_001276761.1 |
| TP53 | 7157 | Asenjo Neurosurgery Institute | hg38 | chr17 | 7674926 | 7674926 | + | Missense_Mutation | SNP | C | C | T | HGG_62 | p.R202H | 0,844660194 | NM_001276761.1 |
| TP53 | 7157 | Asenjo Neurosurgery Institute | hg38 | chr17 | 7674945 | 7674945 | + | Nonsense_Mutation | SNP | G | G | A | HGG_4 | p.R196* | 0,09710897 | NM_001276761.1 |
| TP53 | 7157 | Asenjo Neurosurgery Institute | hg38 | chr17 | 7674962 | 7674962 | + | Missense_Mutation | SNP | G | G | A | HGG_37 | p.P190L | 0,852803738 | NM_001276761.1 |
| TP53 | 7157 | Asenjo Neurosurgery Institute | hg38 | chr17 | 7674966 | 7674966 | + | Frame_Shift_Del | DEL | C | C | - | HGG_48 | p.A189fs | 0,106652587 | NM_001276761.1 |
| TP53 | 7157 | Asenjo Neurosurgery Institute | hg38 | chr17 | 7675077 | 7675077 | + | Missense_Mutation | SNP | G | G | C | HGG_17 | p.H179D | 0,926605505 | NM_001276761.1 |
| TP53 | 7157 | Asenjo Neurosurgery Institute | hg38 | chr17 | 7675088 | 7675088 | + | Missense_Mutation | SNP | C | C | T | HGG_48 | p.R175H | 0,589783282 | NM_001276761.1 |
| TP53 | 7157 | Asenjo Neurosurgery Institute | hg38 | chr17 | 7675094 | 7675094 | + | Missense_Mutation | SNP | A | A | C | HGG_3 | p.V173G | 0,600660066 | NM_001276761.1 |
| TP53 | 7157 | Asenjo Neurosurgery Institute | hg38 | chr17 | 7675206 | 7675206 | + | Missense_Mutation | SNP | G | G | C | HGG_61 | p.Q136E | 0,805340224 | NM_001276761.1 |
| TP53 | 7157 | Asenjo Neurosurgery Institute | hg38 | chr17 | 7675238 | 7675238 | + | Splice_Site | SNP | T | T | A | HGG_5 | | 0,51984127 | NM_001276761.1 |
| TP53 | 7157 | Asenjo Neurosurgery Institute | hg38 | chr17 | 7675995 | 7675995 | + | Splice_Site | SNP | G | G | A | HGG_33 | p.T125M | 0,863354037 | NM_001276761.1 |
| TP53 | 7157 | Asenjo Neurosurgery Institute | hg38 | chr17 | 7676028 | 7676028 | + | Missense_Mutation | SNP | A | A | C | HGG_3 | p.L114W | 0,19047619 | NM_001276761.1 |
| NF1 | 4763 | Asenjo Neurosurgery Institute | hg38 | chr17 | 31338792 | 31338792 | + | Missense_Mutation | SNP | C | C | T | HGG_8 | p.P2282L | 0,066666667 | NM_000267.3 |
| EGFR | 1956 | Asenjo Neurosurgery Institute | hg38 | chr7 | 55143387 | 55143387 | + | Missense_Mutation | SNP | G | G | A | HGG_38 | p.R108K | 0,86035313 | NM_001346897.1 |
| EGFR | 1956 | Asenjo Neurosurgery Institute | hg38 | chr7 | 55151307 | 55151307 | + | Missense_Mutation | SNP | T | T | A | HGG_1 | p.D146E | 0,969676994 | NM_001346897.1 |
| EGFR | 1956 | Asenjo Neurosurgery Institute | hg38 | chr7 | 55154128 | 55154128 | + | Missense_Mutation | SNP | G | G | A | HGG_26 | p.A244T | 0,099056604 | NM_001346897.1 |

| | | | | | | | | | | | | | | | | |
|---|---|---|---|---|---|---|---|---|---|---|---|---|---|---|---|---|
| EGFR | 1956 | Asenjo Neurosurgery Institute | hg38 | chr7 | 55154128 | 55154128 | + | Missense_Mutation | SNP | G | G | A | HGG_32 | p.A244T | 0,94488189 | NM_001346897.1 |
| EGFR | 1956 | Asenjo Neurosurgery Institute | hg38 | chr7 | 55154128 | 55154128 | + | Missense_Mutation | SNP | G | G | A | HGG_35 | p.A244T | 0,194331984 | NM_001346897.1 |
| EGFR | 1956 | Asenjo Neurosurgery Institute | hg38 | chr7 | 55154129 | 55154129 | + | Missense_Mutation | SNP | C | C | T | HGG_6 | p.A244V | 0,909090909 | NM_001346897.1 |
| EGFR | 1956 | Asenjo Neurosurgery Institute | hg38 | chr7 | 55154129 | 55154129 | + | Missense_Mutation | SNP | C | C | T | HGG_20 | p.A244V | 0,403703704 | NM_001346897.1 |
| EGFR | 1956 | Asenjo Neurosurgery Institute | hg38 | chr7 | 55154129 | 55154129 | + | Missense_Mutation | SNP | C | C | T | HGG_42 | p.A244V | 0,090909091 | NM_001346897.1 |
| EGFR | 1956 | Asenjo Neurosurgery Institute | hg38 | chr7 | 55154129 | 55154129 | + | Missense_Mutation | SNP | C | C | A | HGG_53 | p.A244D | 0,175510204 | NM_001346897.1 |
| EGFR | 1956 | Asenjo Neurosurgery Institute | hg38 | chr7 | 55154129 | 55154129 | + | Missense_Mutation | SNP | C | C | T | HGG_60 | p.A244V | 0,751210068 | NM_001346897.1 |
| EGFR | 1956 | Asenjo Neurosurgery Institute | hg38 | chr7 | 55154142 | 55154142 | + | Missense_Mutation | SNP | G | G | C | HGG_32 | p.K248N | 0,94481982 | NM_001346897.1 |
| EGFR | 1956 | Asenjo Neurosurgery Institute | hg38 | chr7 | 55155847 | 55155847 | + | Missense_Mutation | SNP | G | G | A | HGG_51 | p.D258N | 0,82300885 | NM_001346897.1 |
| EGFR | 1956 | Asenjo Neurosurgery Institute | hg38 | chr7 | 55156614 | 55156614 | + | Missense_Mutation | SNP | C | C | T | HGG_4 | p.T318I | 0,395308427 | NM_001346897.1 |
| EGFR | 1956 | Asenjo Neurosurgery Institute | hg38 | chr7 | 55160222 | 55160222 | + | Missense_Mutation | SNP | T | T | C | HGG_62 | p.V416A | 0,379844961 | NM_001346897.1 |
| EGFR | 1956 | Asenjo Neurosurgery Institute | hg38 | chr7 | 55165350 | 55165350 | + | Missense_Mutation | SNP | G | G | T | HGG_1 | p.G553V | 0,807002562 | NM_001346897.1 |
| EGFR | 1956 | Asenjo Neurosurgery Institute | hg38 | chr7 | 55165416 | 55165416 | + | Missense_Mutation | SNP | G | G | A | HGG_68 | p.C575Y | 0,257425743 | NM_001346897.1 |
| EGFR | 1956 | Asenjo Neurosurgery Institute | hg38 | chr7 | 55198792 | 55198792 | + | Missense_Mutation | SNP | T | T | C | HGG_3 | p.I881T | 0,225806452 | NM_001346897.1 |
| EGFR | 1956 | Asenjo Neurosurgery Institute | hg38 | chr7 | 55198792 | 55198792 | + | Missense_Mutation | SNP | T | T | C | HGG_5 | p.I881T | 0,5 | NM_001346897.1 |
| ERN1 | 2081 | Asenjo Neurosurgery Institute | hg38 | chr17 | 64044182 | 64044182 | + | Missense_Mutation | SNP | G | G | T | HGG_22 | p.L914M | 0,4579 | NM_001433.3 |
| ERN1 | 2081 | Asenjo Neurosurgery Institute | hg38 | chr17 | 64044182 | 64044182 | + | Missense_Mutation | SNP | G | G | T | HGG_23 | p.L914M | 1 | NM_001433.3 |
| ERN1 | 2081 | Asenjo Neurosurgery Institute | hg38 | chr17 | 64044182 | 64044182 | + | Missense_Mutation | SNP | G | G | T | HGG_25 | p.L914M | 0,4031 | NM_001433.3 |
| ERN1 | 2081 | Asenjo Neurosurgery Institute | hg38 | chr17 | 64044182 | 64044182 | + | Missense_Mutation | SNP | G | G | T | HGG_41 | p.L914M | 0,0738 | NM_001433.3 |
| ERN1 | 2081 | Asenjo Neurosurgery Institute | hg38 | chr17 | 64044182 | 64044182 | + | Missense_Mutation | SNP | G | G | T | HGG_43 | p.L914M | 0,5333 | NM_001433.3 |
| ERN1 | 2081 | Asenjo Neurosurgery Institute | hg38 | chr17 | 64044182 | 64044182 | + | Missense_Mutation | SNP | G | G | T | HGG_53 | p.L914M | 0,1837 | NM_001433.3 |
| ERN1 | 2081 | Asenjo Neurosurgery Institute | hg38 | chr17 | 64044182 | 64044182 | + | Missense_Mutation | SNP | G | G | T | HGG_58 | p.L914M | 0,496 | NM_001433.3 |
| ERN1 | 2081 | Asenjo Neurosurgery Institute | hg38 | chr17 | 64044182 | 64044182 | + | Missense_Mutation | SNP | G | G | T | HGG_70 | p.L914M | 0,4465 | NM_001433.3 |
| ERN1 | 2081 | Asenjo Neurosurgery Institute | hg38 | chr17 | 64054341 | 64054341 | + | Missense_Mutation | SNP | T | T | C | HGG_55 | p.E621G | 0,121212121 | NM_001433.3 |
| ERN1 | 2081 | Asenjo Neurosurgery Institute | hg38 | chr17 | 64054411 | 64054411 | + | Missense_Mutation | SNP | C | C | T | HGG_33 | p.V598M | 0,053571429 | NM_001433.3 |
| ATRX | 546 | Asenjo Neurosurgery Institute | hg38 | chrX | 77599534 | 77599534 | + | Missense_Mutation | SNP | C | C | T | HGG_8 | p.G1945R | 0,076086957 | NM_000489.4 |
| ATRX | 546 | Asenjo Neurosurgery Institute | hg38 | chrX | 77682178 | 77682178 | + | Missense_Mutation | SNP | A | A | C | HGG_2 | p.C1026W | 0,120171674 | NM_000489.4 |
| ATRX | 546 | Asenjo Neurosurgery Institute | hg38 | chrX | 77682178 | 77682178 | + | Missense_Mutation | SNP | A | A | C | HGG_3 | p.C1026W | 0,060606061 | NM_000489.4 |
| ATRX | 546 | Asenjo Neurosurgery Institute | hg38 | chrX | 77682178 | 77682178 | + | Missense_Mutation | SNP | A | A | C | HGG_5 | p.C1026W | 0,104089219 | NM_000489.4 |
| ATRX | 546 | Asenjo Neurosurgery Institute | hg38 | chrX | 77682178 | 77682178 | + | Missense_Mutation | SNP | A | A | C | HGG_10 | p.C1026W | 0,101769912 | NM_000489.4 |
| ATRX | 546 | Asenjo Neurosurgery Institute | hg38 | chrX | 77684938 | 77684938 | + | Splice_Site | SNP | C | C | T | HGG_55 | | 0,282828283 | NM_000489.4 |
| PTEN | 5728 | Asenjo Neurosurgery Institute | hg38 | chr10 | 87864513 | 87864513 | + | Missense_Mutation | SNP | G | G | A | HGG_8 | p.R15K | 0,73136646 | NM_001304717.2 |
| PTEN | 5728 | Asenjo Neurosurgery Institute | hg38 | chr10 | 87894052 | 87894052 | + | Missense_Mutation | SNP | G | G | T | HGG_64 | p.G36V | 0,3014862 | NM_001304717.2 |
| PTEN | 5728 | Asenjo Neurosurgery Institute | hg38 | chr10 | 87925518 | 87925518 | + | Missense_Mutation | SNP | T | T | C | HGG_26 | p.L57S | 0,361084221 | NM_001304717.2 |
| PTEN | 5728 | Asenjo Neurosurgery Institute | hg38 | chr10 | 87925551 | 87925551 | + | Missense_Mutation | SNP | A | A | G | HGG_66 | p.Y68C | 0,284848485 | NM_001304717.2 |
| PTEN | 5728 | Asenjo Neurosurgery Institute | hg38 | chr10 | 87925553 | 87925553 | + | Missense_Mutation | SNP | A | A | G | HGG_22 | p.N69D | 0,436781609 | NM_001304717.2 |
| PTEN | 5728 | Asenjo Neurosurgery Institute | hg38 | chr10 | 87931089 | 87931089 | + | Splice_Site | SNP | G | G | C | HGG_53 | p.V85L | 0,116564417 | NM_001304717.2 |
| PTEN | 5728 | Asenjo Neurosurgery Institute | hg38 | chr10 | 87933224 | 87933225 | + | Frame_Shift_Ins | INS | - | - | G | HGG_27 | p.E157fs | 0,437238494 | NM_001304717.2 |
| PTEN | 5728 | Asenjo Neurosurgery Institute | hg38 | chr10 | 87933253 | 87933253 | + | Splice_Site | DEL | T | T | - | HGG_29 | | 0,173669468 | NM_001304717.2 |
| PTEN | 5728 | Asenjo Neurosurgery Institute | hg38 | chr10 | 87952235 | 87952235 | + | Missense_Mutation | SNP | C | C | T | HGG_24 | p.P204S | 0,623042506 | NM_001304717.2 |
| PTEN | 5728 | Asenjo Neurosurgery Institute | hg38 | chr10 | 87957970 | 87957970 | + | Missense_Mutation | SNP | G | G | T | HGG_39 | p.G251V | 0,343675418 | NM_001304717.2 |
| PTEN | 5728 | Asenjo Neurosurgery Institute | hg38 | chr10 | 87957976 | 87957976 | + | Missense_Mutation | SNP | T | T | G | HGG_9 | p.I253S | 0,088235294 | NM_001304717.2 |
| PTEN | 5728 | Asenjo Neurosurgery Institute | hg38 | chr10 | 87957976 | 87957976 | + | Missense_Mutation | SNP | T | T | G | HGG_47 | p.I253S | 0,860869565 | NM_001304717.2 |
| IDH1 | 3417 | Asenjo Neurosurgery Institute | hg38 | chr2 | 208248388 | 208248388 | + | Missense_Mutation | SNP | C | C | T | HGG_5 | p.R132H | 0,452879581 | NM_001282387.1 |
| IDH1 | 3417 | Asenjo Neurosurgery Institute | hg38 | chr2 | 208248388 | 208248388 | + | Missense_Mutation | SNP | C | C | T | HGG_13 | p.R132H | 0,378808549 | NM_001282387.1 |
| IDH1 | 3417 | Asenjo Neurosurgery Institute | hg38 | chr2 | 208248388 | 208248388 | + | Missense_Mutation | SNP | C | C | T | HGG_17 | p.R132H | 0,406797116 | NM_001282387.1 |
| IDH1 | 3417 | Asenjo Neurosurgery Institute | hg38 | chr2 | 208248388 | 208248388 | + | Missense_Mutation | SNP | C | C | T | HGG_19 | p.R132H | 0,406189555 | NM_001282387.1 |
| IDH1 | 3417 | Asenjo Neurosurgery Institute | hg38 | chr2 | 208248388 | 208248388 | + | Missense_Mutation | SNP | C | C | T | HGG_28 | p.R132H | 0,434746467 | NM_001282387.1 |
| IDH1 | 3417 | Asenjo Neurosurgery Institute | hg38 | chr2 | 208248388 | 208248388 | + | Missense_Mutation | SNP | C | C | T | HGG_31 | p.R132H | 0,359765833 | NM_001282387.1 |
| IDH1 | 3417 | Asenjo Neurosurgery Institute | hg38 | chr2 | 208248388 | 208248388 | + | Missense_Mutation | SNP | C | C | T | HGG_37 | p.R132H | 0,400977995 | NM_001282387.1 |
| IDH1 | 3417 | Asenjo Neurosurgery Institute | hg38 | chr2 | 208248388 | 208248388 | + | Missense_Mutation | SNP | C | C | T | HGG_50 | p.R132H | 0,444116199 | NM_001282387.1 |
| IDH1 | 3417 | Asenjo Neurosurgery Institute | hg38 | chr2 | 208248389 | 208248389 | + | Missense_Mutation | SNP | G | G | T | HGG_3 | p.R132S | 0,363953488 | NM_001282387.1 |
| H3-3A | | Asenjo Neurosurgery Institute | hg38 | chr1 | 226064434 | 226064434 | + | Missense_Mutation | SNP | A | A | T | HGG_55 | p.K28M | 0,422413793 | NM_002107.4 |

Supplementary Table 3

| Chr | Position | Mean coverage | Median coverage | HGG_1 | HGG_2 | HGG_3 | HGG_4 | HGG_5 | HGG_6 | HGG_7 | HGG_8 | HGG_9 | HGG_10 | HGG_11 | HGG_12 | HGG_13 | HGG_14 | HGG_15 | HGG_16 | HGG_17 | HGG_18 | HGG_19 | HGG_20 | HGG_21 | HGG_22 | HGG_23 | HGG_24 | HGG_25 | HGG_26 | HGG_27 | HGG_28 | HGG_29 | HGG_30 |
|---|---|---|---|---|---|---|---|---|---|---|---|---|---|---|---|---|---|---|---|---|---|---|---|---|---|---|---|---|---|---|---|---|---|
| chr5 | 1295113 | 452,9 | 289,5 | 101 | 114 | 847 | 203 | 691 | 254 | 648 | 420 | 191 | 669 | 306 | 103 | 107 | 447 | 502 | 226 | 163 | 178 | 156 | 221 | 247 | 272 | 168 | 136 | 402 | 484 | 119 | 152 | 167 | 179 |
| chr5 | 1295135 | 864,4 | 584,5 | 208 | 212 | 1622 | 373 | 1283 | 478 | 1239 | 810 | 361 | 1271 | 626 | 189 | 197 | 873 | 958 | 449 | 312 | 329 | 283 | 412 | 481 | 505 | 315 | 255 | 753 | 901 | 235 | 284 | 323 | 357 |
| chr17 | 7669615 | 7974,9 | 4119,5 | 493 | 20932 | 32908 | 2396 | 18484 | 712 | 1566 | 13090 | 5172 | 5620 | 7951 | 591 | 9735 | 14707 | 12396 | 12423 | 7460 | 13346 | 7758 | 996 | 1376 | 2710 | 3288 | 22781 | 2292 | 8164 | 2908 | 5722 | 800 | 753 |
| chr17 | 7673776 | 9149,3 | 4580,5 | 774 | 24509 | 13318 | 2300 | 11863 | 0 | 1161 | 7263 | 5180 | 8369 | 7707 | 552 | 13581 | 16250 | 14192 | 12256 | 8377 | 14468 | 9996 | 1187 | 1183 | 2646 | 3688 | 27255 | 1870 | 8663 | 3844 | 5915 | 1294 | 1132 |
| chr17 | 7673803 | 11343,4 | 5272,5 | 1035 | 33331 | 7158 | 2666 | 6264 | 0 | 1467 | 3850 | 6035 | 12222 | 9806 | 696 | 16610 | 17825 | 19147 | 18199 | 11101 | 19265 | 11908 | 1392 | 1535 | 3469 | 4805 | 35727 | 2257 | 10605 | 4584 | 6869 | 1528 | 1451 |
| chr17 | 7673820 | 9650,4 | 4353,0 | 900 | 28510 | 705 | 2319 | 465 | 0 | 1304 | 432 | 5154 | 10904 | 8572 | 620 | 14223 | 14872 | 17053 | 16795 | 9845 | 16955 | 9860 | 1108 | 1328 | 2942 | 4337 | 31410 | 1819 | 8716 | 4059 | 5943 | 1322 | 1252 |
| chr17 | 7673826 | 8801,8 | 3742,5 | 864 | 27906 | 45 | 1939 | 31 | 2 | 1133 | 10 | 4515 | 10662 | 7788 | 553 | 12799 | 12838 | 15943 | 15983 | 9178 | 15877 | 9072 | 1040 | 1247 | 2895 | 3763 | 29222 | 1769 | 8433 | 3340 | 4933 | 1067 | 1116 |
| chr17 | 7674199 | 1314,8 | 680,5 | 131 | 2590 | 2 | 443 | 0 | 0 | 224 | 1 | 816 | 1145 | 1504 | 39 | 2081 | 3237 | 2088 | 1878 | 977 | 2156 | 1345 | 98 | 113 | 309 | 433 | 4212 | 323 | 1076 | 797 | 1217 | 175 | 106 |
| chr17 | 7674218 | 2439,6 | 872,0 | 111 | 2715 | 212 | 806 | 124 | 64 | 434 | 64 | 1740 | 1020 | 2002 | 74 | 4081 | 5498 | 3042 | 4752 | 3048 | 2746 | 3104 | 494 | 488 | 928 | 1203 | 12275 | 788 | 2702 | 1184 | 2297 | 316 | 88 |
| chr17 | 7674220 | 2440,6 | 872,0 | 111 | 2716 | 212 | 809 | 124 | 64 | 434 | 64 | 1740 | 1020 | 2003 | 74 | 4082 | 5503 | 3043 | 4756 | 3050 | 2746 | 3105 | 494 | 488 | 928 | 1203 | 12279 | 789 | 2706 | 1184 | 2297 | 316 | 88 |
| chr17 | 7674227 | 2441,5 | 872,5 | 111 | 2714 | 212 | 809 | 124 | 64 | 434 | 64 | 1741 | 1020 | 2004 | 74 | 4085 | 5509 | 3043 | 4763 | 3051 | 2748 | 3105 | 496 | 489 | 929 | 1203 | 12286 | 789 | 2706 | 1183 | 2296 | 316 | 88 |
| chr17 | 7674230 | 2442,5 | 872,5 | 111 | 2714 | 212 | 810 | 124 | 64 | 434 | 64 | 1741 | 1020 | 2004 | 74 | 4087 | 5512 | 3045 | 4765 | 3052 | 2748 | 3107 | 496 | 489 | 929 | 1204 | 12293 | 790 | 2707 | 1183 | 2297 | 316 | 88 |
| chr17 | 7674233 | 2442,5 | 872,0 | 111 | 2715 | 212 | 810 | 124 | 64 | 434 | 64 | 1741 | 1020 | 2004 | 74 | 4087 | 5511 | 3047 | 4765 | 3052 | 2748 | 3107 | 496 | 488 | 928 | 1204 | 12294 | 790 | 2708 | 1182 | 2297 | 316 | 88 |
| chr17 | 7674893 | 9811,4 | 4406,0 | 829 | 28018 | 26171 | 2317 | 14405 | 0 | 1165 | 12737 | 4680 | 11151 | 6912 | 651 | 12296 | 15242 | 14887 | 11783 | 9323 | 12524 | 8799 | 1500 | 1531 | 2903 | 4034 | 34645 | 2064 | 9864 | 1959 | 5683 | 1133 | 1174 |
| chr17 | 7674894 | 9811,3 | 4406,5 | 829 | 28018 | 26167 | 2317 | 14404 | 0 | 1165 | 12737 | 4681 | 11151 | 6912 | 651 | 12296 | 15242 | 14887 | 11782 | 9322 | 12524 | 8799 | 1500 | 1530 | 2902 | 4034 | 34645 | 2064 | 9862 | 1959 | 5683 | 1133 | 1174 |
| chr17 | 7674905 | 10538,2 | 4573,5 | 1006 | 32466 | 13233 | 2369 | 7318 | 0 | 1239 | 6450 | 4878 | 13225 | 7710 | 670 | 14537 | 17734 | 17778 | 15988 | 10652 | 15598 | 10003 | 1510 | 1644 | 3266 | 4292 | 38951 | 2129 | 11142 | 1474 | 5788 | 1224 | 1245 |
| chr17 | 7674926 | 8066,0 | 3046,0 | 841 | 26443 | 3 | 1725 | 1 | 0 | 937 | 0 | 3616 | 10775 | 6005 | 491 | 12027 | 14396 | 14759 | 14349 | 8639 | 13122 | 7943 | 1067 | 1250 | 2573 | 3388 | 31263 | 1537 | 8726 | 2309 | 4051 | 904 | 933 |
| chr17 | 7674945 | 8141,0 | 3085,5 | 842 | 26589 | 10 | 1723 | 3 | 3 | 947 | 0 | 3655 | 10850 | 6050 | 498 | 12108 | 14477 | 14834 | 14430 | 8691 | 13173 | 7986 | 1072 | 1256 | 2582 | 3462 | 31650 | 1544 | 8776 | 2342 | 4140 | 915 | 932 |
| chr17 | 7674962 | 8471,7 | 3218,0 | 821 | 26417 | 204 | 1949 | 58 | 52 | 1025 | 31 | 4041 | 10766 | 6299 | 522 | 12834 | 15041 | 14985 | 15350 | 9097 | 13381 | 8725 | 1227 | 1381 | 2803 | 3634 | 33561 | 1737 | 9439 | 2538 | 4746 | 969 | 920 |
| chr17 | 7674966 | 4733,5 | 1773,0 | 423 | 13877 | 204 | 1177 | 58 | 52 | 613 | 31 | 2413 | 5559 | 3569 | 275 | 7523 | 8927 | 8259 | 8719 | 5045 | 7405 | 5171 | 692 | 705 | 1553 | 1935 | 18905 | 1005 | 5270 | 1611 | 3083 | 560 | 430 |
| chr17 | 7675077 | 3418,7 | 1736,5 | 165 | 10338 | 13418 | 983 | 7836 | 279 | 562 | 5527 | 2029 | 2304 | 2872 | 188 | 5167 | 7564 | 5140 | 4668 | 3513 | 5692 | 3650 | 519 | 520 | 1001 | 1173 | 11492 | 879 | 3359 | 1439 | 2543 | 424 | 242 |
| chr17 | 7675088 | 3420,4 | 1737,0 | 166 | 10345 | 13433 | 983 | 7841 | 278 | 562 | 5531 | 2030 | 2304 | 2872 | 188 | 5166 | 7565 | 5147 | 4668 | 3519 | 5693 | 3656 | 519 | 520 | 1000 | 1171 | 11495 | 879 | 3359 | 1439 | 2543 | 424 | 242 |
| chr17 | 7675094 | 3067,5 | 1550,5 | 196 | 8542 | 19395 | 878 | 9748 | 8 | 466 | 6818 | 2010 | 2594 | 2405 | 147 | 3810 | 6825 | 3712 | 2221 | 2656 | 3614 | 2712 | 329 | 280 | 781 | 784 | 8437 | 731 | 2727 | 1254 | 2266 | 383 | 267 |
| chr17 | 7675206 | 7301,8 | 2583,0 | 378 | 15351 | 1252 | 2460 | 269 | 236 | 1528 | 236 | 4478 | 4385 | 5505 | 546 | 13465 | 15306 | 10707 | 13948 | 8387 | 9698 | 9204 | 1536 | 1724 | 2519 | 3280 | 31231 | 2087 | 9140 | 2647 | 6731 | 799 | 456 |
| chr17 | 7675238 | 3880,6 | 1067,5 | 58 | 4439 | 1227 | 1583 | 259 | 230 | 1013 | 224 | 2610 | 814 | 2705 | 319 | 8009 | 9402 | 4953 | 8079 | 5226 | 4190 | 5885 | 974 | 1010 | 1599 | 1817 | 18080 | 1316 | 5492 | 1539 | 4491 | 401 | 102 |
| chr17 | 7675995 | 6353,6 | 3085,0 | 734 | 16992 | 19732 | 1522 | 16286 | 34 | 811 | 14310 | 3317 | 8808 | 4434 | 485 | 6984 | 9356 | 7583 | 8041 | 7046 | 6710 | 5213 | 937 | 1044 | 2052 | 2644 | 21044 | 1115 | 5660 | 2014 | 3222 | 804 | 816 |
| chr17 | 7676028 | 10406,7 | 4500,5 | 1127 | 28723 | 22296 | 2274 | 25940 | 2 | 1122 | 22990 | 4901 | 13496 | 6797 | 732 | 12299 | 16623 | 14513 | 14085 | 10965 | 11682 | 8966 | 1823 | 1923 | 3478 | 4572 | 36348 | 1870 | 10003 | 2609 | 4429 | 1175 | 1392 |
| chr17 | 31338792 | 2538,7 | 540,5 | 42 | 3339 | 731 | 476 | 638 | 140 | 133 | 254 | 1111 | 881 | 1392 | 297 | 3006 | 3619 | 2750 | 4082 | 3557 | 1755 | 3403 | 299 | 214 | 1775 | 2678 | 10138 | 489 | 3399 | 1403 | 1400 | 263 | 52 |
| chr7 | 55143387 | 17426,6 | 9522,0 | 19647 | 22010 | 24953 | 12054 | 17327 | 1 | 16468 | 15565 | 3692 | 12480 | 7343 | 1754 | 15868 | 20500 | 22612 | 16302 | 8172 | 15612 | 10209 | 20642 | 27046 | 5831 | 149893 | 34770 | 1634 | 127874 | 6271 | 4688 | 7753 | 16099 |
| chr7 | 55151307 | 29209,4 | 15681,5 | 30740 | 30782 | 50000 | 17230 | 36190 | 4 | 30270 | 47966 | 5636 | 24513 | 10269 | 3837 | 20269 | 27981 | 33750 | 22420 | 14983 | 19856 | 11434 | 50975 | 89906 | 9452 | 210455 | 67023 | 3523 | 222711 | 5441 | 5728 | 11494 | 29899 |
| chr7 | 55154128 | 12946,0 | 8955,5 | 16253 | 20951 | 26174 | 16211 | 15255 | 23 | 31349 | 12836 | 4766 | 7563 | 9401 | 1221 | 13455 | 25996 | 18797 | 12396 | 6528 | 14978 | 7529 | 18777 | 20974 | 3809 | 65399 | 29140 | 2110 | 13540 | 6206 | 4272 | 8811 | 9341 |
| chr7 | 55154129 | 12946,3 | 8953,0 | 16253 | 20954 | 26174 | 16208 | 15255 | 23 | 31348 | 12835 | 4765 | 7564 | 9402 | 1221 | 13457 | 25993 | 18797 | 12396 | 6527 | 14977 | 7529 | 18781 | 20980 | 3811 | 65399 | 29140 | 2110 | 13539 | 6205 | 4270 | 8808 | 9341 |
| chr7 | 55154142 | 12929,2 | 8947,5 | 16054 | 20794 | 25884 | 16211 | 15101 | 23 | 31348 | 12754 | 4766 | 7475 | 9397 | 1221 | 13454 | 25982 | 18784 | 12395 | 6526 | 14973 | 7529 | 18786 | 20978 | 3808 | 65300 | 29121 | 2108 | 13532 | 6196 | 4265 | 8797 | 9331 |
| chr7 | 55155847 | 7383,0 | 4358,0 | 8309 | 11907 | 1 | 7036 | 2 | 1 | 12985 | 3 | 2024 | 5394 | 4894 | 954 | 11038 | 16730 | 13845 | 10840 | 4847 | 10458 | 4435 | 11784 | 29997 | 2662 | 40128 | 16780 | 1028 | 13053 | 2007 | 1765 | 4350 | 7606 |
| chr7 | 55156614 | 15004,4 | 6102,0 | 1830 | 6280 | 1259 | 17484 | 860 | 11466 | 30124 | 724 | 5517 | 1383 | 8149 | 2425 | 20336 | 19456 | 16042 | 22885 | 10090 | 11624 | 12027 | 39106 | 66755 | 6529 | 116720 | 45216 | 2521 | 29931 | 6512 | 6408 | 9083 | 4840 |
| chr7 | 55160222 | 21037,8 | 11071,0 | 13635 | 20180 | 16915 | 12588 | 23420 | 5575 | 19791 | 26272 | 4615 | 12273 | 9153 | 2944 | 20365 | 25013 | 27309 | 24350 | 11023 | 19903 | 12027 | 28101 | 61745 | 7853 | 204495 | 48524 | 2119 | 32349 | 7401 | 6658 | 12972 | 21667 |
| chr7 | 55165350 | 20282,0 | 11814,5 | 10607 | 20016 | 38002 | 30357 | 23779 | 9173 | 36707 | 23787 | 5687 | 7796 | 11011 | 2113 | 18854 | 25443 | 23393 | 16316 | 8252 | 18440 | 9333 | 31330 | 69029 | 5054 | 112994 | 37834 | 2979 | 147682 | 6586 | 6434 | 14500 | 15509 |
| chr7 | 55165416 | 11986,6 | 5461,5 | 10403 | 13268 | 197 | 16181 | 118 | 0 | 18545 | 105 | 2200 | 7000 | 5503 | 1248 | 11957 | 15362 | 15196 | 10905 | 4864 | 11109 | 4994 | 12654 | 41533 | 2534 | 95961 | 24750 | 1057 | 79789 | 3679 | 3046 | 7408 | 12828 |
| chr7 | 55191792 | 19280,5 | 11365,0 | 20040 | 17258 | 11068 | 26041 | 5862 | 8 | 34532 | 6001 | 4359 | 12222 | 10218 | 38596 | 15990 | 20962 | 23397 | 19810 | 8598 | 15813 | 7103 | 26561 | 71423 | 4914 | 132494 | 35160 | 2088 | 119159 | 4789 | 3787 | 12900 | 17880 |
| chr17 | 64044182 | 278,6 | 195,5 | 234 | 585 | 207 | 410 | 256 | 580 | 180 | 248 | 209 | 378 | 368 | 58 | 201 | 157 | 500 | 535 | 214 | 249 | 167 | 335 | 290 | 387 | 249 | 207 | 219 | 507 | 164 | 194 | 176 | 192 |
| chr17 | 64054341 | 339,3 | 209,5 | 262 | 151 | 402 | 95 | 204 | 125 | 153 | 123 | 141 | 215 | 303 | 57 | 95 | 241 | 301 | 92 | 281 | 230 | 104 | 313 | 209 | 323 | 155 | 229 | 277 | 710 | 102 | 130 | 160 | 208 |
| chr17 | 64054411 | 323,0 | 197,5 | 247 | 146 | 378 | 90 | 185 | 120 | 146 | 103 | 128 | 209 | 287 | 54 | 95 | 211 | 277 | 94 | 265 | 208 | 94 | 308 | 194 | 305 | 143 | 227 | 256 | 635 | 94 | 114 | 147 | 200 |
| chrX | 77599534 | 2203,1 | 592,0 | 35 | 3658 | 551 | 228 | 327 | 135 | 171 | 98 | 1156 | 993 | 2005 | 198 | 2235 | 8974 | 2054 | 5898 | 3361 | 3936 | 6198 | 546 | 769 | 1535 | 3882 | 5691 | 1215 | 7865 | 1243 | 3803 | 74 | 100 |
| chrX | 77682178 | 29,6 | 4,5 | 6 | 249 | 352 | 0 | 287 | 3 | 0 | 121 | 3 | 255 | 14 | 1 | 12 | 46 | 20 | 26 | 16 | 38 | 32 | 1 | 2 | 12 | 4 | 22 | 9 | 51 | 3 | 10 | 0 | 0 |
| chrX | 77682909 | 5158,5 | 1391,5 | 40 | 5227 | 911 | 965 | 833 | 350 | 402 | 290 | 2558 | 1479 | 6113 | 408 | 5550 | 18564 | 4555 | 13964 | 6000 | 9582 | 11985 | 1265 | 1432 | 2955 | 9528 | 10246 | 2126 | 13771 | 3221 | 6491 | 296 | 232 |
| chrX | 77684938 | 7143,4 | 2826,0 | 92 | 16910 | 20505 | 727 | 14453 | 1018 | 377 | 7135 | 2532 | 5715 | 7270 | 593 | 6272 | 22253 | 8831 | 19154 | 6347 | 20793 | 12547 | 1612 | 2793 | 3683 | 10739 | 14475 | 1990 | 14985 | 2859 | 4826 | 487 | 1349 |
| chr10 | 87864488 | 6017,6 | 1842,0 | 135 | 13505 | 22498 | 1550 | 4488 | 601 | 589 | 4682 | 1924 | 1746 | 3554 | 362 | 1760 | 11400 | 9162 | 10287 | 7022 | 8154 | 8035 | 349 | 350 | 2379 | 4504 | 12217 | 1356 | 6736 | 2193 | 201 | 541 | 479 |
| chr10 | 87864513 | 7862,0 | 2022,0 | 188 | 21975 | 43438 | 1615 | 8655 | 981 | 584 | 9259 | 2054 | 2806 | 4443 | 431 | 1867 | 12222 | 12770 | 12226 | 7812 | 11500 | 8533 | 446 | 438 | 2726 | 5555 | 13853 | 1473 | 7461 | 2191 | 236 | 708 | 812 |
| chr10 | 87894052 | 12967,5 | 1620,0 | 68 | 6153 | 1130 | 1434 | 581 | 280 | 378 | 446 | 4385 | 2552 | 5343 | 1073 | 2702 | 9436 | 13152 | 24932 | 24939 | 9961 | 18605 | 931 | 766 | 7621 | 19044 | 36804 | 2771 | 17340 | 6156 | 677 | 1121 | 494 |
| chr10 | 87925518 | 7268,6 | 1227,5 | 54 | 5133 | 3615 | 576 | 1282 | 179 | 170 | 1188 | 2419 | 1267 | 2791 | 307 | 5129 | 5227 | 4489 | 12187 | 11864 | 5182 | 12774 | 335 | 752 | 5031 | 6779 | 22515 | 2219 | 12203 | 5601 | 604 | 451 | 180 |
| chr10 | 87925551 | 15445,5 | 3534,0 | 192 | 23198 | 35281 | 883 | 13984 | 514 | 267 | 12755 | 4065 | 5842 | 5042 | 566 | 8144 | 9311 | 11451 | 22609 | 22811 | 11506 | 20941 | 551 | 1328 | 10752 | 11184 | 45533 | 4325 | 21446 | 9480 | 930 | 923 | 698 |
| chr10 | 87925553 | 15436,6 | 3534,0 | 192 | 23197 | 35284 | 883 | 13984 | 514 | 267 | 12755 | 4065 | 5838 | 5041 | 565 | 8141 | 9307 | 11443 | 22588 | 22797 | 11499 | 20925 | 551 | 1327 | 10742 | 11164 | 45502 | 4325 | 21429 | 9476 | 930 | 923 | 698 |
| chr10 | 87931089 | 6110,1 | 1888,0 | 102 | 16357 | 31998 | 627 | 12381 | 537 | 202 | 10584 | 1966 | 4008 | 2948 | 300 | 5852 | 8027 | 5908 | 10445 | 6838 | 9950 | 10431 | 170 | 456 | 2889 | 4351 | 10891 | 1242 | 7069 | 4897 | 494 | 572 | 635 |
| chr10 | 87933225 | 2750,9 | 436,0 | 25 | 1334 | 255 | 654 | 164 | 101 | 141 | 94 | 1297 | 475 | 1385 | 193 | 2096 | 3020 | 1523 | 4961 | 4178 | 1823 | 3662 | 145 | 333 | 1019 | 4565 | 7857 | 627 | 2347 | 3144 | 263 | 375 | 89 |
| chr10 | 87933253 | 3012,6 | 627,0 | 23 | 1753 | 1329 | 694 | 494 | 107 | 162 | 416 | 1379 | 521 | 1564 | 216 | 2432 | 3865 | 1834 | 5473 | 4466 | 2448 | 4164 | 148 | 355 | 1063 | 4767 | 8292 | 670 | 2517 | 3503 | 282 | 318 | 108 |
| chr10 | 87952235 | 12643,0 | 4741,5 | 404 | 28333 | 25215 | 1946 | 27214 | 2 | 372 | 24593 | 3926 | 10595 | 5668 | 868 | 10051 | 13474 | 10886 | 16714 | 16348 | 13856 | 15114 | 458 | 1024 | 5589 | 10471 | 27191 | 2458 | 14441 | 7245 | 441 | 1397 | 2103 |
| chr10 | 87957970 | 17860,4 | 5430,5 | 436 | 31836 | 486 | 3170 | 374 | 138 | 794 | 131 | 7185 | 14323 | 11100 | 1122 | 15616 | 21554 | 16947 | 28741 | 26476 | 21815 | 22582 | 750 | 2159 | 8420 | 15583 | 41705 | 5149 | 23376 | 11650 | 1153 | 2094 | 2386 |
| chr10 | 87957976 | 21464,4 | 5464,5 | 467 | 33757 | 897 | 3205 | 383 | 139 | 804 | 243 | 7253 | 15124 | 11176 | 1407 | 19343 | 26797 | 20060 | 36091 | 34347 | 25157 | 29511 | 1024 | 2894 | 11016 | 20310 | 54003 | 5198 | 29955 | 11754 | 1163 | 2105 | 2518 |
| chr2 | 208248388 | 12617,2 | 5378,5 | 744 | 31150 | 16146 | 2216 | 36787 | 310 | 1012 | 21627 | 5858 | 12510 | 8594 | 1245 | 15055 | 13558 | 16820 | 13443 | 13855 | 17258 | 14000 | 2455 | 1568 | 6673 | 8884 | 31630 | 2318 | 15750 | 4434 | 6190 | 1668 | 2633 |
| chr2 | 208248389 | 12609,5 | 5374,5 | 744 | 31150 | 16147 | 2216 | 36502 | 310 | 1012 | 21627 | 5860 | 12510 | 8595 | 1245 | 15020 | 13558 | 16820 | 13443 | 13810 | 17257 | 13972 | 2455 | 1568 | 6673 | 8884 | 31630 | 2318 | 15750 | 4436 | 6160 | 1668 | 2633 |
| chr1 | 226064434 | 5657,1 | 3750,0 | 812 | 801 | 4528 | 318 | 3150 | 832 | 2192 | 7694 | 1993 | 595 | 3683 | 1793 | 163 | 1995 | 5552 | 390 | 1236 | 2762 | 1312 | 2109 | 1330 | 1874 | 1808 | 1616 | 3817 | 7226 | 1407 | 1393 | 2861 | 4469 |

| Chr | Position | HGG_31 | HGG_32 | HGG_33 | HGG_34 | HGG_35 | HGG_36 | HGG_37 | HGG_38 | HGG_39 | HGG_40 | HGG_41 | HGG_42 | HGG_43 | HGG_44 | HGG_45 | HGG_46 | HGG_47 | HGG_48 | HGG_49 | HGG_50 | HGG_51 | HGG_52 | HGG_53 | HGG_54 | HGG_55 | HGG_56 | HGG_57 | HGG_58 | HGG_59 | HGG_60 | HGG_61 | HGG_62 | HGG_63 | HGG_64 | HGG_65 | HGG_66 | HGG_67 | HGG_68 | HGG_69 | HGG_70 |
|---|---|---|---|---|---|---|---|---|---|---|---|---|---|---|---|---|---|---|---|---|---|---|---|---|---|---|---|---|---|---|---|---|---|---|---|---|---|---|---|---|---|
| chr5 | 1295113 | 93 | 273 | 66 | 617 | 213 | 172 | 471 | 466 | 167 | 326 | 922 | 1136 | 148 | 73 | 515 | 2102 | 484 | 1661 | 491 | 695 | 354 | 1817 | 56 | 209 | 90 | 2178 | 622 | 503 | 510 | 964 | 394 | 167 | 95 | 437 | 649 | 142 | 169 | 748 | 627 | 1011 |
| chr5 | 1295135 | 179 | 549 | 131 | 1203 | 386 | 332 | 872 | 880 | 310 | 620 | 1785 | 2177 | 300 | 130 | 978 | 3952 | 931 | 3218 | 936 | 1322 | 697 | 3475 | 102 | 416 | 178 | 4135 | 1153 | 989 | 1008 | 1831 | 794 | 307 | 177 | 836 | 1212 | 293 | 342 | 1420 | 1204 | 1852 |
| chr17 | 7669615 | 8349 | 63 | 2254 | 4887 | 881 | 148 | 4237 | 638 | 3753 | 4002 | 16993 | 464 | 9923 | 28 | 86 | 35199 | 1603 | 1770 | 1165 | 32891 | 660 | 42427 | 652 | 570 | 10896 | 15329 | 24725 | 241 | 256 | 19186 | 18192 | 17192 | 2474 | 11983 | 4623 | 13070 | 5806 | 509 | 415 | 171 |
| chr17 | 7673776 | 13124 | 63 | 3563 | 4938 | 1233 | 166 | 4695 | 816 | 3999 | 8494 | 16997 | 259 | 13777 | 20 | 65 | 39614 | 1434 | 1846 | 3047 | 38071 | 707 | 46077 | 600 | 744 | 13794 | 18404 | 32970 | 271 | 246 | 33644 | 21724 | 29639 | 3512 | 17791 | 4466 | 19546 | 7535 | 702 | 806 | 261 |
| chr17 | 7673803 | 15747 | 91 | 4801 | 5522 | 1549 | 230 | 5848 | 1046 | 4950 | 10310 | 21011 | 229 | 17816 | 23 | 90 | 53912 | 2101 | 2191 | 4023 | 52158 | 835 | 60175 | 724 | 1039 | 16641 | 23864 | 42659 | 394 | 337 | 44965 | 28589 | 31016 | 5023 | 22548 | 5523 | 21789 | 9284 | 821 | 1017 | 342 |
| chr17 | 7673820 | 13678 | 77 | 4308 | 4695 | 1352 | 206 | 5031 | 915 | 4369 | 8870 | 18578 | 174 | 15272 | 19 | 85 | 48397 | 1898 | 1926 | 3437 | 46359 | 724 | 53362 | 602 | 926 | 13581 | 20117 | 36905 | 362 | 302 | 39320 | 24391 | 24490 | 4447 | 20230 | 4868 | 17671 | 7977 | 673 | 917 | 290 |
| chr17 | 7673826 | 11100 | 69 | 3948 | 3696 | 1202 | 171 | 4488 | 760 | 3722 | 7808 | 15752 | 123 | 14335 | 15 | 75 | 44316 | 1811 | 1577 | 3273 | 43752 | 616 | 48849 | 577 | 859 | 12763 | 19183 | 34584 | 327 | 279 | 37124 | 23179 | 20363 | 4249 | 17013 | 4101 | 15367 | 7148 | 594 | 757 | 236 |
| chr17 | 7674199 | 1741 | 14 | 539 | 863 | 160 | 29 | 708 | 107 | 709 | 1558 | 3431 | 27 | 1433 | 4 | 4 | 4999 | 137 | 404 | 399 | 5849 | 170 | 6548 | 108 | 69 | 1586 | 1871 | 4885 | 56 | 39 | 3448 | 4095 | 5156 | 653 | 4238 | 843 | 4196 | 1196 | 75 | 134 | 38 |
| chr17 | 7674218 | 1481 | 14 | 384 | 1720 | 191 | 61 | 675 | 71 | 1619 | 1136 | 5730 | 60 | 4046 | 4 | 18 | 14228 | 526 | 712 | 217 | 14449 | 258 | 19096 | 174 | 258 | 3587 | 4766 | 13218 | 82 | 106 | 2399 | 2671 | 5699 | 397 | 7093 | 1639 | 2490 | 816 | 52 | 153 | 77 |
| chr17 | 7674220 | 1481 | 14 | 383 | 1722 | 191 | 61 | 675 | 71 | 1619 | 1136 | 5733 | 60 | 4047 | 4 | 18 | 14232 | 526 | 712 | 217 | 14458 | 258 | 19104 | 174 | 258 | 3587 | 4770 | 13223 | 82 | 106 | 2399 | 2671 | 5699 | 397 | 7097 | 1640 | 2491 | 816 | 52 | 153 | 77 |
| chr17 | 7674227 | 1481 | 14 | 383 | 1721 | 191 | 61 | 677 | 71 | 1620 | 1137 | 5734 | 60 | 4048 | 4 | 18 | 14243 | 526 | 712 | 217 | 14462 | 258 | 19112 | 174 | 260 | 3588 | 4769 | 13237 | 82 | 106 | 2393 | 2667 | 5699 | 397 | 7095 | 1641 | 2490 | 816 | 52 | 153 | 77 |
| chr17 | 7674230 | 1481 | 14 | 383 | 1721 | 191 | 61 | 678 | 71 | 1620 | 1137 | 5737 | 60 | 4050 | 4 | 18 | 14254 | 526 | 712 | 217 | 14472 | 258 | 19117 | 175 | 260 | 3589 | 4772 | 13240 | 82 | 106 | 2396 | 2667 | 5702 | 398 | 7094 | 1641 | 2490 | 816 | 52 | 153 | 77 |
| chr17 | 7674233 | 1481 | 14 | 383 | 1721 | 191 | 61 | 678 | 71 | 1619 | 1137 | 5738 | 60 | 4051 | 4 | 18 | 14254 | 526 | 711 | 217 | 14473 | 258 | 19117 | 175 | 260 | 3590 | 4774 | 13238 | 82 | 106 | 2396 | 2666 | 5702 | 396 | 7097 | 1641 | 2488 | 816 | 52 | 153 | 77 |
| chr17 | 7674893 | 11281 | 49 | 2877 | 4948 | 1421 | 137 | 4685 | 944 | 4098 | 8795 | 15600 | 298 | 15195 | 24 | 81 | 49720 | 1191 | 1723 | 3179 | 44075 | 747 | 55014 | 625 | 930 | 11848 | 18606 | 40021 | 289 | 356 | 33611 | 24404 | 23573 | 3481 | 14621 | 4132 | 14971 | 7385 | 578 | 674 | 234 |
| chr17 | 7674894 | 11280 | 49 | 2877 | 4948 | 1421 | 137 | 4685 | 944 | 4098 | 8795 | 15596 | 298 | 15195 | 24 | 81 | 49723 | 1191 | 1723 | 3179 | 44075 | 747 | 55013 | 625 | 931 | 11848 | 18605 | 40021 | 289 | 356 | 33611 | 24405 | 23573 | 3481 | 14621 | 4132 | 14971 | 7385 | 578 | 674 | 233 |
| chr17 | 7674905 | 11417 | 56 | 3314 | 5068 | 1459 | 147 | 4855 | 905 | 4255 | 9293 | 16669 | 235 | 17116 | 23 | 92 | 55565 | 1447 | 1807 | 3369 | 52272 | 793 | 60933 | 623 | 1020 | 12618 | 19717 | 45916 | 332 | 353 | 39560 | 26773 | 23421 | 3909 | 17079 | 4236 | 16373 | 7518 | 573 | 784 | 257 |
| chr17 | 7674926 | 8890 | 48 | 2729 | 3617 | 1086 | 117 | 3741 | 659 | 3311 | 7172 | 12604 | 118 | 13544 | 18 | 64 | 45673 | 1198 | 1440 | 2555 | 43245 | 605 | 48454 | 418 | 784 | 9346 | 14668 | 36830 | 261 | 256 | 31365 | 20031 | 16496 | 3004 | 13715 | 3088 | 12610 | 5546 | 407 | 623 | 210 |
| chr17 | 7674945 | 9023 | 48 | 2738 | 3664 | 1093 | 121 | 3762 | 667 | 3398 | 7238 | 12881 | 123 | 13648 | 18 | 64 | 46343 | 1209 | 1488 | 2563 | 43625 | 606 | 49092 | 427 | 786 | 9418 | 14756 | 37154 | 263 | 258 | 31430 | 20101 | 16716 | 3019 | 13915 | 3152 | 12674 | 5596 | 407 | 629 | 214 |
| chr17 | 7674962 | 8558 | 50 | 2588 | 3980 | 1094 | 135 | 3706 | 654 | 3557 | 7044 | 13794 | 141 | 14520 | 18 | 70 | 47904 | 1386 | 1545 | 2487 | 46463 | 635 | 51804 | 509 | 831 | 10221 | 15645 | 39663 | 281 | 266 | 30837 | 19755 | 17835 | 2963 | 15279 | 3473 | 12127 | 5407 | 391 | 634 | 215 |
| chr17 | 7674966 | 4565 | 25 | 1367 | 2390 | 539 | 83 | 1963 | 311 | 2130 | 3715 | 8554 | 82 | 8074 | 9 | 39 | 26241 | 803 | 881 | 1268 | 25867 | 367 | 28850 | 305 | 449 | 5794 | 8786 | 22409 | 152 | 143 | 15773 | 10301 | 10579 | 1536 | 9631 | 2153 | 6606 | 2859 | 204 | 367 | 124 |
| chr17 | 7675077 | 2724 | 17 | 698 | 1966 | 267 | 48 | 1758 | 154 | 1511 | 2559 | 6106 | 158 | 4515 | 13 | 23 | 13676 | 509 | 670 | 998 | 12941 | 281 | 17618 | 226 | 268 | 3957 | 4921 | 13102 | 97 | 74 | 7905 | 7292 | 8958 | 854 | 6282 | 1715 | 6352 | 2135 | 207 | 196 | 73 |
| chr17 | 7675088 | 2722 | 17 | 697 | 1964 | 267 | 48 | 1758 | 154 | 1509 | 2560 | 6111 | 158 | 4513 | 13 | 23 | 13674 | 509 | 669 | 1002 | 12950 | 281 | 17636 | 225 | 268 | 3965 | 4929 | 13103 | 97 | 74 | 7908 | 7299 | 8962 | 856 | 6289 | 1716 | 6357 | 2134 | 207 | 196 | 73 |
| chr17 | 7675094 | 3852 | 8 | 831 | 1750 | 352 | 31 | 1641 | 222 | 1376 | 2874 | 5660 | 134 | 3186 | 4 | 16 | 9979 | 246 | 547 | 857 | 9256 | 311 | 13391 | 227 | 148 | 3371 | 3874 | 9400 | 77 | 57 | 7389 | 6506 | 12177 | 1052 | 5249 | 1460 | 7353 | 2357 | 238 | 188 | 81 |
| chr17 | 7675206 | 4835 | 64 | 1265 | 5110 | 780 | 145 | 2363 | 390 | 3848 | 4278 | 22196 | 245 | 12340 | 21 | 61 | 45431 | 1704 | 2119 | 1196 | 40290 | 630 | 51985 | 622 | 699 | 9552 | 17560 | 36192 | 234 | 318 | 11318 | 6226 | 13727 | 1368 | 15791 | 5287 | 6460 | 4073 | 229 | 481 | 201 |
| chr17 | 7675238 | 1384 | 35 | 200 | 3255 | 285 | 82 | 571 | 36 | 2385 | 881 | 14878 | 160 | 6558 | 11 | 41 | 25453 | 1021 | 1405 | 4 | 23767 | 380 | 30363 | 395 | 353 | 5472 | 9993 | 22203 | 100 | 166 | 1000 | 1114 | 5396 | 153 | 8940 | 3323 | 933 | 548 | 30 | 220 | 99 |
| chr17 | 7675995 | 7690 | 36 | 1407 | 3371 | 693 | 121 | 3318 | 438 | 2948 | 4494 | 9108 | 252 | 9717 | 19 | 54 | 33313 | 642 | 1078 | 1917 | 25082 | 449 | 42312 | 219 | 627 | 6827 | 7209 | 19640 | 147 | 212 | 28035 | 12688 | 13451 | 2716 | 8903 | 2708 | 8334 | 3830 | 399 | 437 | 162 |
| chr17 | 7676028 | 12218 | 46 | 2379 | 4812 | 1126 | 146 | 5298 | 745 | 4008 | 7593 | 12505 | 373 | 16908 | 35 | 100 | 57943 | 1031 | 1440 | 3493 | 45551 | 712 | 69783 | 362 | 1124 | 12040 | 12728 | 34736 | 197 | 366 | 45986 | 19331 | 22431 | 4184 | 12936 | 3986 | 12952 | 6162 | 672 | 645 | 229 |
| chr17 | 31338792 | 861 | 5 | 64 | 1201 | 86 | 75 | 174 | 5 | 856 | 73 | 2200 | 38 | 2762 | 2 | 19 | 35877 | 706 | 344 | 0 | 34575 | 86 | 19073 | 64 | 180 | 1485 | 1866 | 12360 | 69 | 36 | 1110 | 189 | 1090 | 207 | 5203 | 592 | 115 | 68 | 5 | 206 | 67 |
| chr7 | 55143387 | 12052 | 1217 | 1912 | 4647 | 7916 | 3078 | 2706 | 7956 | 3484 | 3155 | 12527 | 2648 | 17751 | 428 | 2341 | 62327 | 1488 | 2659 | 2421 | 55842 | 2142 | 58095 | 7122 | 14257 | 11178 | 10496 | 34267 | 6599 | 3252 | 99479 | 12612 | 23729 | 8835 | 18656 | 2530 | 14501 | 3944 | 3703 | 7392 | 3445 |
| chr7 | 55151307 | 19749 | 1970 | 2626 | 7549 | 17229 | 4017 | 5872 | 19833 | 5507 | 6627 | 16224 | 6073 | 31525 | 997 | 5058 | 133503 | 3137 | 3821 | 4890 | 66487 | 4376 | 119772 | 9411 | 33120 | 16248 | 15265 | 53751 | 9143 | 7705 | 137306 | 22614 | 28410 | 11770 | 18922 | 4128 | 16098 | 9216 | 6741 | 8409 | 4821 |
| chr7 | 55154128 | 10111 | 948 | 2101 | 4982 | 8558 | 2374 | 4190 | 9100 | 3849 | 4859 | 16643 | 2949 | 12829 | 283 | 1826 | 42296 | 1482 | 3276 | 2519 | 29914 | 2592 | 46542 | 8207 | 9406 | 11270 | 9909 | 25718 | 6880 | 3313 | 62049 | 14789 | 28461 | 4802 | 20456 | 3859 | 18861 | 6900 | 3909 | 6353 | 2793 |
| chr7 | 55154129 | 10110 | 948 | 2101 | 4981 | 8557 | 2374 | 4187 | 9098 | 3849 | 4855 | 16645 | 2948 | 12831 | 283 | 1826 | 42297 | 1484 | 3276 | 2518 | 29917 | 2591 | 46541 | 8210 | 9410 | 11276 | 9911 | 25720 | 6879 | 3313 | 62061 | 14791 | 28460 | 4803 | 20456 | 3858 | 18862 | 6900 | 3909 | 6353 | 2792 |
| chr7 | 55154142 | 10108 | 945 | 2100 | 4978 | 8554 | 2369 | 4187 | 9098 | 3848 | 4861 | 16646 | 2945 | 12831 | 283 | 1823 | 42281 | 1484 | 3277 | 2524 | 29905 | 2592 | 46510 | 8212 | 9409 | 11286 | 9916 | 25720 | 6881 | 3310 | 62090 | 14795 | 28463 | 4783 | 20454 | 3860 | 18865 | 6902 | 3911 | 6355 | 2793 |
| chr7 | 55155847 | 4366 | 475 | 1220 | 1932 | 4924 | 1143 | 2450 | 4774 | 1762 | 2566 | 6646 | 1742 | 7685 | 171 | 1249 | 29690 | 979 | 1429 | 1386 | 21544 | 5874 | 28555 | 3289 | 6677 | 3382 | 4238 | 14440 | 3339 | 1812 | 43208 | 6390 | 10306 | 2390 | 8636 | 1464 | 7839 | 3644 | 2577 | 2540 | 1215 |
| chr7 | 55156614 | 5114 | 1229 | 504 | 5742 | 5924 | 3237 | 1723 | 2261 | 5092 | 1393 | 19291 | 5686 | 21050 | 425 | 4970 | 86565 | 5219 | 4229 | 31 | 66878 | 6514 | 82860 | 12097 | 16123 | 10258 | 13892 | 44162 | 5353 | 4034 | 12977 | 1968 | 18754 | 390 | 24271 | 3755 | 2593 | 1348 | 1083 | 5085 | 2646 |
| chr7 | 55160222 | 14687 | 1453 | 2138 | 6974 | 9951 | 3746 | 4108 | 11119 | 4478 | 3617 | 16156 | 7697 | 22233 | 460 | 3709 | 102635 | 2385 | 2935 | 2623 | 77144 | 6417 | 90647 | 7573 | 18277 | 9714 | 10584 | 44451 | 7214 | 3720 | 128200 | 15171 | 23408 | 6385 | 20068 | 3087 | 14946 | 4602 | 4742 | 7962 | 3993 |
| chr7 | 55165350 | 12937 | 1593 | 2222 | 7288 | 11293 | 185 | 4753 | 13129 | 4771 | 5888 | 21414 | 9312 | 18393 | 515 | 4562 | 65562 | 2543 | 4543 | 3031 | 45105 | 8054 | 70563 | 12612 | 16024 | 12336 | 12762 | 32049 | 7909 | 4612 | 84459 | 20174 | 30904 | 5420 | 22205 | 4515 | 21138 | 8555 | 6297 | 7370 | 3748 |
| chr7 | 55165416 | 7105 | 1283 | 1418 | 3150 | 7700 | 152 | 2052 | 9335 | 2576 | 2978 | 12136 | 2208 | 10354 | 433 | 2080 | 51226 | 1265 | 2336 | 1714 | 37134 | 5326 | 44099 | 5250 | 10275 | 5420 | 7664 | 23387 | 8547 | 3409 | 64191 | 10140 | 13105 | 3335 | 14443 | 2251 | 10303 | 3965 | 2749 | 5033 | 3141 |
| chr7 | 55191792 | 12453 | 1974 | 2723 | 5065 | 16131 | 4252 | 4492 | 14665 | 4198 | 5199 | 16504 | 5241 | 18218 | 723 | 4388 | 77693 | 3057 | 3190 | 3236 | 45818 | 8267 | 69967 | 8338 | 17308 | 11662 | 14490 | 32165 | 10860 | 5195 | 88284 | 15858 | 20535 | 7607 | 18346 | 3587 | 13856 | 7298 | 4574 | 8692 | 4495 |
| chr17 | 64044182 | 35 | 271 | 169 | 153 | 148 | 148 | 72 | 186 | 110 | 134 | 151 | 186 | 86 | 69 | 207 | 2537 | 743 | 332 | 471 | 173 | 170 | 301 | 57 | 75 | 42 | 632 | 197 | 142 | 199 | 177 | 137 | 152 | 122 | 230 | 108 | 64 | 140 | 183 | 508 | 827 |
| chr17 | 64054341 | 79 | 416 | 128 | 325 | 74 | 293 | 94 | 179 | 85 | 317 | 361 | 358 | 166 | 119 | 157 | 3055 | 841 | 1242 | 505 | 210 | 169 | 1049 | 114 | 125 | 74 | 1387 | 226 | 237 | 384 | 287 | 182 | 175 | 57 | 279 | 191 | 126 | 227 | 328 | 491 | 2246 |
| chr17 | 64054411 | 75 | 410 | 124 | 303 | 70 | 273 | 90 | 176 | 76 | 306 | 359 | 333 | 160 | 107 | 147 | 2949 | 791 | 1232 | 465 | 195 | 155 | 1034 | 107 | 120 | 70 | 1380 | 220 | 229 | 367 | 270 | 177 | 174 | 56 | 269 | 183 | 122 | 213 | 318 | 466 | 2081 |
| chrX | 77599534 | 581 | 12 | 352 | 1586 | 143 | 49 | 202 | 18 | 835 | 186 | 5180 | 24 | 4022 | 10 | 14 | 4152 | 897 | 659 | 0 | 11038 | 196 | 19380 | 66 | 295 | 5230 | 4146 | 18285 | 29 | 103 | 436 | 603 | 1803 | 98 | 2338 | 1781 | 177 | 281 | 6 | 46 | 24 |
| chrX | 77682178 | 5 | 0 | 4 | 6 | 1 | 0 | 6 | 0 | 3 | 0 | 14 | 0 | 31 | 0 | 0 | 73 | 0 | 1 | 2 | 45 | 0 | 95 | 0 | 1 | 18 | 24 | 56 | 0 | 0 | 19 | 24 | 4 | 25 | 7 | 3 | 4 | 7 | 0 | 0 | 0 |
| chrX | 77682909 | 1892 | 59 | 542 | 3564 | 266 | 219 | 426 | 42 | 2415 | 383 | 8473 | 141 | 8806 | 9 | 60 | 37319 | 3373 | 1804 | 2 | 33660 | 441 | 41899 | 168 | 641 | 9568 | 7859 | 35368 | 79 | 163 | 1213 | 1351 | 5563 | 192 | 6974 | 3399 | 517 | 500 | 14 | 278 | 116 |
| chrX | 77684938 | 3161 | 74 | 2093 | 3292 | 393 | 186 | 1901 | 299 | 1433 | 1632 | 4266 | 250 | 13214 | 32 | 64 | 45133 | 3792 | 1058 | 1400 | 32325 | 412 | 53860 | 143 | 1048 | 7669 | 4089 | 42227 | 101 | 283 | 10914 | 10634 | 2894 | 1125 | 7145 | 2149 | 3343 | 2041 | 198 | 356 | 89 |
| chr10 | 87864488 | 2898 | 19 | 595 | 2081 | 414 | 106 | 2182 | 123 | 1982 | 974 | 8503 | 71 | 8753 | 22 | 30 | 69132 | 1259 | 1058 | 804 | 59569 | 225 | 31769 | 237 | 576 | 9356 | 7946 | 33086 | 93 | 111 | 10090 | 4290 | 5058 | 594 | 7361 | 2832 | 3250 | 445 | 85 | 411 | 81 |
| chr10 | 87864513 | 4300 | 21 | 992 | 2408 | 517 | 122 | 3479 | 206 | 1990 | 1556 | 8979 | 64 | 11739 | 35 | 26 | 86362 | 1494 | 1079 | 1575 | 73984 | 256 | 38716 | 257 | 767 | 11128 | 10049 | 38737 | 98 | 141 | 18507 | 7956 | 4504 | 1020 | 7619 | 2948 | 5920 | 804 | 148 | 492 | 87 |
| chr10 | 87894052 | 4445 | 73 | 603 | 5046 | 762 | 289 | 1806 | 38 | 5197 | 510 | 7787 | 178 | 24341 | 34 | 74 | 215544 | 3738 | 1252 | 12 | 171155 | 484 | 90935 | 315 | 2045 | 18904 | 18305 | 75005 | 158 | 401 | 7000 | 546 | 6183 | 508 | 16752 | 3963 | 848 | 98 | 55 | 789 | 273 |
| chr10 | 87925518 | 2055 | 4 | 330 | 3625 | 304 | 113 | 755 | 35 | 2525 | 189 | 10835 | 64 | 12915 | 12 | 49 | 98742 | 1115 | 616 | 103 | 91964 | 140 | 45656 | 197 | 595 | 13998 | 15866 | 50429 | 74 | 141 | 4467 | 1668 | 2263 | 428 | 10385 | 2801 | 342 | 89 | 31 | 300 | 81 |
| chr10 | 87925551 | 5022 | 12 | 1144 | 6216 | 540 | 171 | 2360 | 137 | 4185 | 651 | 19226 | 162 | 27267 | 29 | 68 | 200044 | 1743 | 873 | 2163 | 167550 | 227 | 94872 | 306 | 1104 | 26523 | 28460 | 97506 | 117 | 250 | 34304 | 12386 | 3003 | 1839 | 16966 | 4490 | 1511 | 400 | 131 | 573 | 145 |
| chr10 | 87925553 | 5022 | 12 | 1144 | 6214 | 540 | 171 | 2358 | 137 | 4185 | 651 | 19226 | 162 | 27257 | 29 | 68 | 199893 | 1743 | 871 | 2163 | 167416 | 227 | 94826 | 306 | 1102 | 26511 | 28437 | 97436 | 117 | 250 | 34295 | 12384 | 3003 | 1839 | 16967 | 4490 | 1511 | 400 | 131 | 573 | 145 |
| chr10 | 87931089 | 4016 | 2 | 1127 | 2944 | 239 | 86 | 2180 | 119 | 1602 | 912 | 6835 | 158 | 8039 | 7 | 46 | 54088 | 825 | 589 | 1058 | 65285 | 147 | 24565 | 195 | 376 | 6141 | 6593 | 23342 | 49 | 76 | 15159 | 6224 | 3485 | 1810 | 9057 | 1769 | 4429 | 425 | 145 | 364 | 69 |
| chr10 | 87933225 | 1782 | 11 | 155 | 1987 | 182 | 91 | 397 | 16 | 1781 | 171 | 4977 | 39 | 4177 | 15 | 43 | 42062 | 1096 | 496 | 2 | 29732 | 103 | 19178 | 45 | 358 | 3926 | 3774 | 15068 | 63 | 79 | 998 | 367 | 1801 | 125 | 7310 | 1355 | 257 | 45 | 8 | 272 | 75 |
| chr10 | 87933253 | 2098 | 10 | 224 | 2150 | 197 | 94 | 503 | 23 | 1872 | 213 | 5636 | 44 | 4446 | 16 | 46 | 44291 | 1132 | 560 | 42 | 31795 | 111 | 20086 | 47 | 371 | 4195 | 3993 | 16223 | 63 | 85 | 1635 | 655 | 2034 | 170 | 7951 | 1454 | 599 | 61 | 11 | 305 | 78 |
| chr10 | 87952235 | 25739 | 28 | 3385 | 5142 | 1288 | 194 | 5513 | 479 | 4896 | 2669 | 10135 | 227 | 17744 | 14 | 77 | 129352 | 1169 | 1391 | 3088 | 109893 | 373 | 62958 | 351 | 1194 | 14025 | 11152 | 52560 | 197 | 193 | 57959 | 13168 | 13670 | 4587 | 14992 | 2845 | 10780 | 1114 | 450 | 1389 | 232 |
| chr10 | 87957970 | 26237 | 26 | 4858 | 8491 | 1925 | 440 | 7420 | 604 | 8878 | 3550 | 24994 | 253 | 27159 | 50 | 207 | 219764 | 4129 | 2945 | 4020 | 174414 | 745 | 97233 | 560 | 1976 | 25529 | 27555 | 80604 | 382 | 499 | 60376 | 17632 | 15969 | 5712 | 30275 | 7525 | 11178 | 4142 | 488 | 1846 | 384 |
| chr10 | 87957976 | 26348 | 26 | 4867 | 8577 | 1933 | 440 | 7448 | 608 | 8942 | 3564 | 25227 | 256 | 34948 | 50 | 212 | 284943 | 4173 | 2968 | 4022 | 224476 | 754 | 127351 | 564 | 2568 | 25824 | 27815 | 106884 | 386 | 504 | 61938 | 17668 | 16133 | 5731 | 30547 | 7606 | 11220 | 4154 | 488 | 1859 | 388 |
| chr2 | 208248388 | 20113 | 572 | 4709 | 6050 | 1980 | 587 | 5373 | 1282 | 5384 | 4841 | 10483 | 1269 | 21242 | 153 | 688 | 101648 | 3732 | 2610 | 3692 | 77388 | 954 | 69113 | 643 | 1527 | 13706 | 11711 | 42295 | 1035 | 754 | 50314 | 19228 | 23910 | 5111 | 16963 | 2996 | 13856 | 4373 | 905 | 1933 | 1689 |
| chr2 | 208248389 | 20072 | 572 | 4709 | 6050 | 1982 | 587 | 5365 | 1283 | 5384 | 4841 | 10481 | 1270 | 21242 | 154 | 688 | 101652 | 3740 | 2610 | 3693 | 77302 | 955 | 69108 | 643 | 1527 | 13705 | 11711 | 42295 | 1035 | 754 | 50312 | 19228 | 23909 | 5111 | 16964 | 2996 | 13857 | 4373 | 905 | 1935 | 1691 |
| chr1 | 226064434 | 8033 | 5270 | 3462 | 11321 | 2415 | 2501 | 2356 | 9031 | 2628 | 5274 | 9640 | 18661 | 4395 | 869 | 7720 | 37477 | 14643 | 15367 | 4426 | 10283 | 11796 | 11291 | 845 | 5791 | 2221 | 14441 | 5185 | 5801 | 4959 | 4566 | 16933 | 6023 | 1212 | 7946 | 3350 | 2585 | 6681 | 12756 | 8496 | 15634 |
